# Peripheral Blood Gene Expression at 3 to 24 Hours Correlates with and Predicts 90-Day Outcome Following Human Ischemic Stroke

**DOI:** 10.1101/2022.06.16.22276291

**Authors:** Hajar Amini, Bodie Knepp, Fernando Rodriguez, Glen C Jickling, Heather Hull, Paulina Carmona-Mora, Cheryl Bushnell, Bradley P Ander, Frank R Sharp, Boryana Stamova

## Abstract

This study identified early immune gene responses in peripheral blood associated with 90-day ischemic stroke (IS) outcomes and an early gene profile that predicted 90-day outcomes. Peripheral blood from the CLEAR trial IS patients was compared to vascular risk factor matched controls. Whole-transcriptome analyses identified genes and networks associated with 90-day IS outcome (NIHSS-NIH Stroke Scale, mRS-modified Rankin Scale). The expression of 467, 526, and 571 genes measured at ≤3, 5 and 24 hours after IS, respectively, were associated with poor 90-day mRS outcome (mRS=3-6), while 49, 100 and 35 associated with good mRS 90-day outcome (mRS=0-2). Poor outcomes were associated with up-regulated *MMP9*, *S100A12*, interleukin-related and STAT3 pathways. Weighted Gene Co-Expression Network Analysis (WGCNA) revealed modules significantly associated with 90-day outcome. Poor outcome modules were enriched in down-regulated T cell and monocyte-specific genes plus up-regulated neutrophil genes and good outcome modules were associated with erythroblasts and megakaryocytes. Using the difference in gene expression between 3 and 24 hours, 10 genes correctly predicted 100% of patients with Good 90-day mRS outcome and 67% with Poor mRS outcome (AUC=0.88) in a validation set. The predictors included *AVPR1A*, which mediates platelet aggregation, release of coagulation factors and exacerbates the brain inflammatory response; and *KCNK1* (*TWIK-1*), a member of a two-pore potassium channel family, which like other potassium channels likely modulates stroke outcomes. This study suggests the immune response after stroke impacts long-term functional outcomes. Furthermore, early post-stroke gene expression may predict stroke outcomes and outcome-associated genes could be targets for improving outcomes.

## Introduction

Changes in gene expression after ischemic stroke (IS) can potentially be used as biomarkers for causes of IS and predicting IS outcome [1–3]. Finding genes associated with long-term recovery after IS will improve our understanding of the pathways involved in recovery mechanisms, and may guide the search for treatment targets and early predictors of IS outcome [2,4–8].

Genetic risk factors have been associated with IS outcome [9], including *PTGIS*, *TBXAS1*, *IL6*, *BDNF*, *CYPC19*, *GPIIIa*, *P2RY1*, *ITGB3*, *PATJ*, *ADAM23*, *GRIA1*, *PARK2*, *ABCB5* and various cytochrome P450 genes [10–13]. Moreover, several clinical variables have been associated with long-term IS outcome including blood pressure [14], glucose levels/diabetes [15, 16], atrial fibrillation [17], and hyperlipidemia [18] in addition to age [19] and sex [11]. Other variables shown to be associated with outcome include stroke severity [20], type of treatment [21], severe complications [22] and stroke etiology [23].

Predicting functional outcome in stroke is challenging partly because of the complexity of the condition and lack of validated prognostic models. Clinical and demographic variables only explain a portion of the variance in long-term IS outcome. Thus, it is important to identify additional biomarkers to explain the remaining long-term outcome variance and to design accurate predictive models. Thus, we have studied the peripheral blood transcriptome of patients after IS to discover genes and pathways that associate with 90-day outcome and use them to develop a molecularly based, pilot machine learning model to predict long-term functional outcome after IS.

## Materials and Methods

### Study Subjects

Peripheral blood was drawn from IS patients at ≤3, 5, and 24 hours (n=36 subjects, 108 samples) as part of the Combined Approach to Lysis Utilizing Eptifibatide and Recombinant Tissue-Type Plasminogen Activator (CLEAR) trial (NCT00250991 at www.Clinical-Trials.gov) [24]. IS subjects were treated with recombinant tissue plasminogen activator (rt-PA) with or without eptifibatide after the within 3h blood sample was obtained. After treatment, blood samples were drawn at 5 hours and 24 hours post-stroke onset. Control subjects included Vascular Risk Factor Control (VRFC) subjects with at least one cardiovascular risk factor (hypertension, diabetes mellitus, hyperlipidemia) recruited from the Sex Age and Variation in Vascular functionalitY (SAVVY, Cheryl Bushnell PI) study (NCT00681681) (n=18) [25]. The IRB at each site approved the study, and each patient or a proxy provided informed consent.

### Sample Processing and Data Analysis

Whole blood was collected into PAXgene tubes (PreAnalytiX) and RNA processed as previously described [4]. Each RNA sample was processed and hybridized on Affymetrix Human U133 Plus 2.0 GeneChips (Affymetrix, Santa Clara, CA). Differences in demographic data between groups were analyzed using a two-tailed *t*-test and χ^2^ analysis where appropriate with *P* <0.05 considered significant. Raw probe-level gene expression values imported into Partek Genomics Suite software (Partek Inc, St Louis, MO) were summarized to probe set-level using Median Polish summarization, and normalized using robust multichip averaging (RMA) and our internal-gene normalization approach [4, 26].

The gene expression at ≤3h, 5h, and 24h was associated with 90-day mRS outcome (modified Rankin Score, categorical variable), and the NIHSS (NIH Stroke Scale, continuous variable). The mRS, subjects with 90-day mRS scores of 0, 1, and 2 were binned into a Good Outcome group (n=26 subjects, 78 samples), and subjects with 90-day mRS of 3, 4, and 5 into a Poor Outcome group (n=10 subjects, 30 samples). This mRS variable is referred to as Binned mRS hereafter. No subject had the maximum mRS=6 (deceased) at 90 days in this dataset.

#### Gene Expression associated with 90-day Binned mRS

An ANCOVA identified genes whose expression significantly associated with 90-day Good and Poor Outcomes (Binned mRS) at each time point (≤3h, 5h, and 24h) after IS compared to VRFC. The ANCOVA model for each time-point was Y*_i_* = μ + Diagnosis (Poor Outcome, Good Outcome, VRFC) + Hypercholesterolemia + Hypertension + Diabetes + Age + Sex + ε*_i_*, where Y*_i_* is gene expression at ≤ 3h, 5h or 24h, μ is the common effect for the whole experiment, and ε*_i_* is the random error. Age was a continuous variable, and Sex and vascular risk factors (Hypercholesterolemia, Hypertension and Diabetes) were considered as binary variables (Yes or No). A false discovery rate (FDR) corrected *P* < 0.05 and a fold change (FC) >∣2∣ were considered significant. We used a cut-off of (FC) >∣1.3∣ and *P* < 0.05 when comparing IS patients with Poor 90-day mRS outcome vs. IS patients with Good mRS outcome.

#### Gene Expression Associated with 90-day NIHSS

Separate analyses identified genes significantly correlated with 90-day NIHSS outcome using gene expression at ≤3h, 5h and 24h. *P* <0.005 was considered significant. The ANCOVA model for each time-point was Y*_i_* = μ + baseline NIHSS + 24hNIHSS + 5dNIHSS + 90dNIHSS + Hypercholesterolemia + Hypertension + Diabetes + Group + Age + Sex + ε*_i_*. Group was either tPA or combined treatment of tPA and eptifibatide. Baseline NIHSS is NIHSS at first draw (within 3h of stroke onset); 24hNIHSS, 5dNIHSS and 90dNIHSS are the NIHSS at 24h, 5 days and 90 days, respectively.

### Weighted Gene Co-Expression Network Construction and Analysis

Networks were generated using the Weighted Gene Co-Expression Network Analysis (WGCNA) package [27]. Separate weighted gene co-expression networks were generated for ≤3, 5, and 24h gene expression following the methods in our recent studies [28]. The details of the analysis for this study are provided in the Supplementary Methods (WGCNA-1).

### Identifying IS Outcome-Associated Modules

Module-outcome associations for Good and Poor outcomes were determined using ANCOVA models in Partek Genomics Suite using the module’s eigengene values. The details of these methods are provided in Supplementary Methods (WGCNA-2).

### Network Visualization and Hub Gene Identification

The v*isantPrepOverall* R function within WGCNA generated a list of intramodular gene connections with parameters numint=10,000 and signed=TRUE [29, 30]. These connections were then imported into Cytoscape for network visualization [31, 32]. Nodes represent genes within the module and edges the connections between genes. Minimum weight cut-off for edges was adjusted for each network to generate a figure with a visually distinguishable number of nodes and connections.

### Cell-Specific Gene Involvement

To identify enrichment in blood cell type-specific genes, differentially expressed gene lists and module gene lists were overlapped with lists of blood cell type-specific genes [33, 34]. The significance of list overlaps was assessed using hypergeometric probability testing (R function *phyper*; *P* < 0.05 considered significant).

### Pathway and Gene Ontology Analyses

Ingenuity Pathway Analysis (IPA^®^, QIAGEN) was performed on all probe set lists as previously described [35] with *P* < 0.05 being considered significant. Details of the Pathway and Gene Ontology Analyses are provided in the Supplementary Methods (Pathway Analyses).

### Predicting 90-day Outcome from Changes in Gene Expression from **≤**3h to 24h after IS

To identify early genes to predict long-term IS outcome, we engineered a new variable for each probe set by calculating the change in gene expression between 24h and 3h post IS. Subjects were divided into a training set (n=25) and a validation set (n=11). Because of the small sample size, we undertook a two-prong approach. First, genes were excluded which upon 1-way ANOVAs were significant for Age, Sex, Hypertension, Diabetes, Hyperlipidemia, and/or Treatment Group. After excluding probe sets significant at *P*<0.05 for any of these variables, ANOVA was performed (Y = μ + 24hNIHSS + Binned_mRS + ε) on the remaining 30,565 probe sets. Probe sets with *P*<0.005 for Poor vs. Good mRS outcome were considered significant. Second, we overlapped the findings from the training set of 25 subjects with the ones from the entire set of 36 and found 10 overlapping probe sets. The 10 probe sets (features) were input into logistic regression and support vector machines (SVM) models with parameters varied as implemented in the scikit-learn package [36]. The classifier was generated from the training set and the best predictive model was deployed on the validation set. The validation set was used to evaluate the performance of the predictors by calculating the sensitivity, specificity, and Receiver Operating Characteristic (ROC) Area Under the Curve (AUC).

## Results

### Subject Demographics

There were no statistically significant differences in age, sex, race, and vascular risk factors between IS subjects and vascular risk factor controls (VRFC) (*P*<0.05, Table 1). The median NIHSS was 10.5, 7.5, 6, 4, and 2 for ≤ 3h, 5h, 24h, 5 days and 90 days post IS. The median mRS at 90d was 2 (Q1=1, Q3=3, range: (0-5)). 26 subjects had good 90-day mRS outcome (0-2), and 10 had Poor 90-day outcome (3-5). No subject had a 90-day mRS of 6 (deceased). Of two subjects with symptomatic hemorrhagic infarction at 24h by CT brain scan, one had a good outcome and one bad.

**Table 1.**
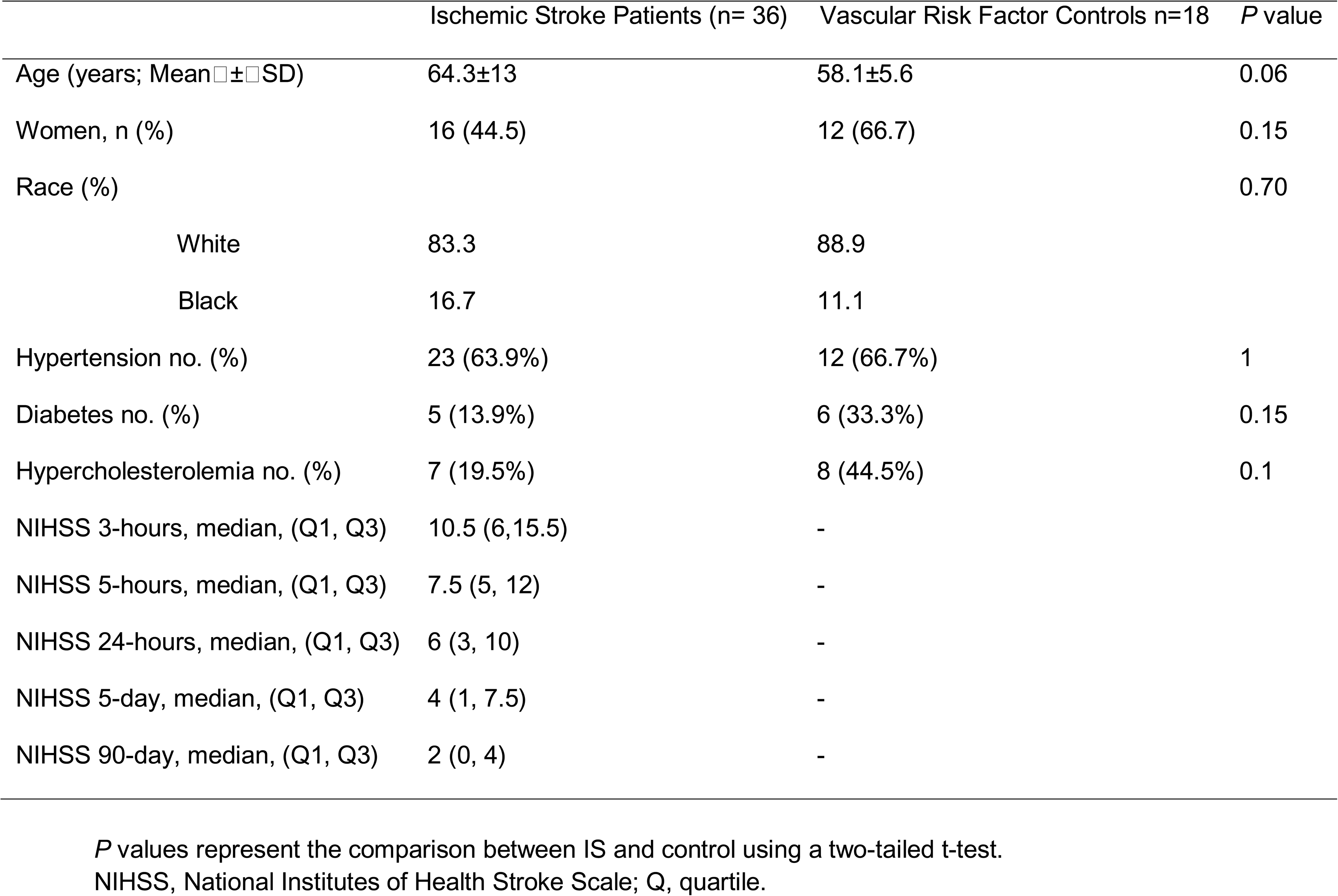
Demographic and clinical characteristics of ischemic stroke (IS) patients and Vascular Risk Factor Controls (Control) subjects.

|  | Ischemic Stroke Patients (n= 36) | Vascular Risk Factor Controls n=18 | *P* value |
| --- | --- | --- | --- |
| Age (years; Mean ± SD) | 64.3±13 | 58.1±5.6 | 0.06 |
| Women, n (%) | 16 (44.5) | 12 (66.7) | 0.15 |
| Race (%) |  |  | 0.70 |
| White | 83.3 | 88.9 |  |
| Black | 16.7 | 11.1 |  |
| Hypertension no. (%) | 23 (63.9%) | 12 (66.7%) | 1 |
| Diabetes no. (%) | 5 (13.9%) | 6 (33.3%) | 0.15 |
| Hypercholesterolemia no. (%) | 7 (19.5%) | 8 (44.5%) | 0.1 |
| NIHSS 3-hours, median, (Q1, Q3) | 10.5 (6,15.5) | - |  |
| NIHSS 5-hours, median, (Q1, Q3) | 7.5 (5, 12) | - |  |
| NIHSS 24-hours, median, (Q1, Q3) | 6 (3, 10) | - |  |
| NIHSS 5-day, median, (Q1, Q3) | 4 (1, 7.5) | - |  |
| NIHSS 90-day, median, (Q1, Q3) | 2 (0, 4) | - |  |

*P* values represent the comparison between IS and control using a two-tailed t-test. NIHSS, National Institutes of Health Stroke Scale; Q, quartile.

### Association of Gene Expression **≤**3h of Stroke with 90-day mRS IS Outcome

#### 3h-Gene Expression Associated with Poor 90-day Functional Outcome (mRS)

Six hundred forty-four probe sets (representing 467 genes) were differentially expressed at ≤3h in subjects with poor 90-day outcome compared to VRFC (FDR-corrected *P*<0.05, fold change (FC)> |2|) (Figure 1a). Of these, 409 probe sets were upregulated and 235 down-regulated (Figure 1a, Table S1A). The 644 probe sets were overrepresented in 47 pathways. Top activated pathways included p38 MAPK, IL-6, IL-1 and STAT3. LXR/RXR was suppressed (Figure 2a, Table S2A). Top over-represented GO terms included B cell receptor signaling, phagocytosis, and immunoglobulin receptor binding, including Immunoglobin Heavy Constant genes such as *IGHG1*, *IGHG*3, *IGHA1*, *IGHA*2, *IGHD*, *IGHM*, and IGHV3-23 (FDR < 0.05) (Table S3A). There was a significant enrichment with neutrophil-specific genes (63/467 genes (13.5%), *P*(overlap)LJ<LJ1E-16) and T cell-specific genes (12/467 genes (2.6%), *P*(overlap)LJ=LJ0.007) (Figure 3a). Most neutrophil-specific genes (60/63) were up-regulated and T cell-specific genes down-regulated in subjects with poor 90-day outcomes.

**Fig. 1.**
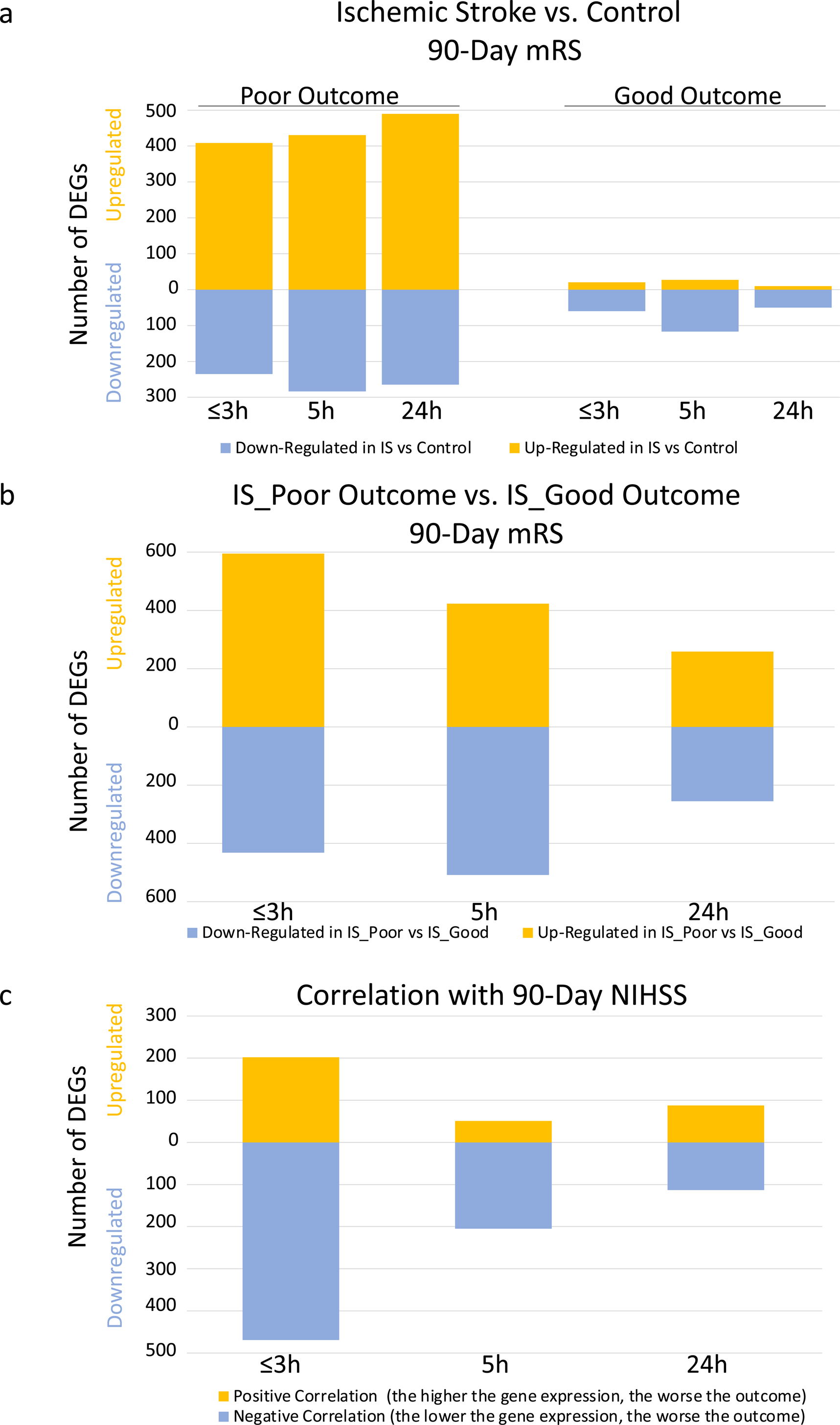
Numbers of up- and down-regulated Differentially Expressed Genes (DEGs) across the three time points ≤3 hours, 5 hours and 24 hours after ischemic stroke. (a) Ischemic Stroke (IS) was compared Vascular Risk Factor Controls (Control) for the Poor Outcome subjects (mRS of 3-5 at 90d) and for the Good Outcome subjects (mRS of 0-2 at 90d). (b) IS Poor Outcome subjects were compared to the IS Good Outcome subjects. (c) Numbers of DEGs that correlated with the 90d NIHSS. The genes from the list for (a) passed FDR-corrected *P* value <0.05 and a fold change (FC)> |2|. The list for the DEGs in (b) had a *P* value <0.05 and a fold change (FC)> |1.3|. The list of the DEGs in (c) had *P* value <0.05. Yellow represents numbers of up-regulated DEGs. Blue represents numbers of down-regulated DEGs. IS – ischemic stroke; mRS – modified Rankin score; NIHSS – National Institutes of Health Stroke Scale.

**Fig. 2.**
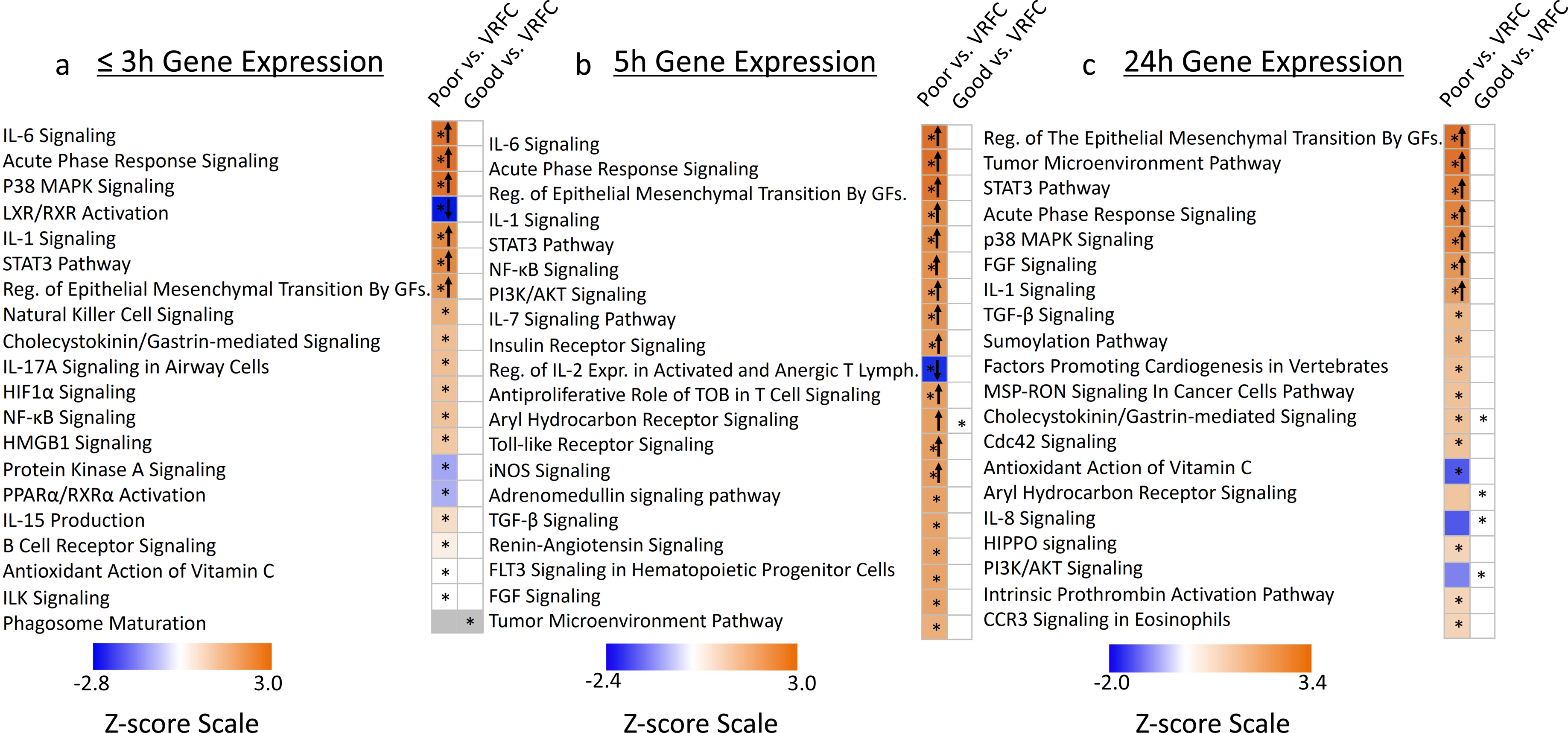
Pathway enrichment for Differentially Expressed Genes (DEGs) for Poor Outcomes (mRS 3-5) compared to Vascular Risk Factor Controls (Poor vs. VRFC) and for Good Outcomes (mRS 0-2) compared to Vascular Risk Factor Controls (Good vs. VRFC). The top 20 most significant activation or suppression relevant pathways for these two comparisons are shown for the three time points after stroke: (a) ≤3h, (b) 5h and (c) and 24h. Blue bars indicate pathway suppression (negative Z-score), and orange indicates activation (positive Z-score), with darker colors representing larger |Z-score|. ↑ (up arrow) represents Z ≥ 2 significant activation in the poor or good 90d mRS IS outcome compared to VRF controls. ↓ (down arrow) represents Z ≤ -2, significant suppression in the poor or good 90d mRS IS outcome compared to VRF controls. The asterisk * represents significantly enriched pathway (*P*<0.05). White cells represent non-significant functions and/or activity pattern prediction Z=0. Grey represents no activity pattern available for the pathway in the IPA knowledge base. Reg. – Regulation; GFs. – Growth Factors; Expr. – Expression; Lymph. – Lymphocytes.

**Fig. 3.**
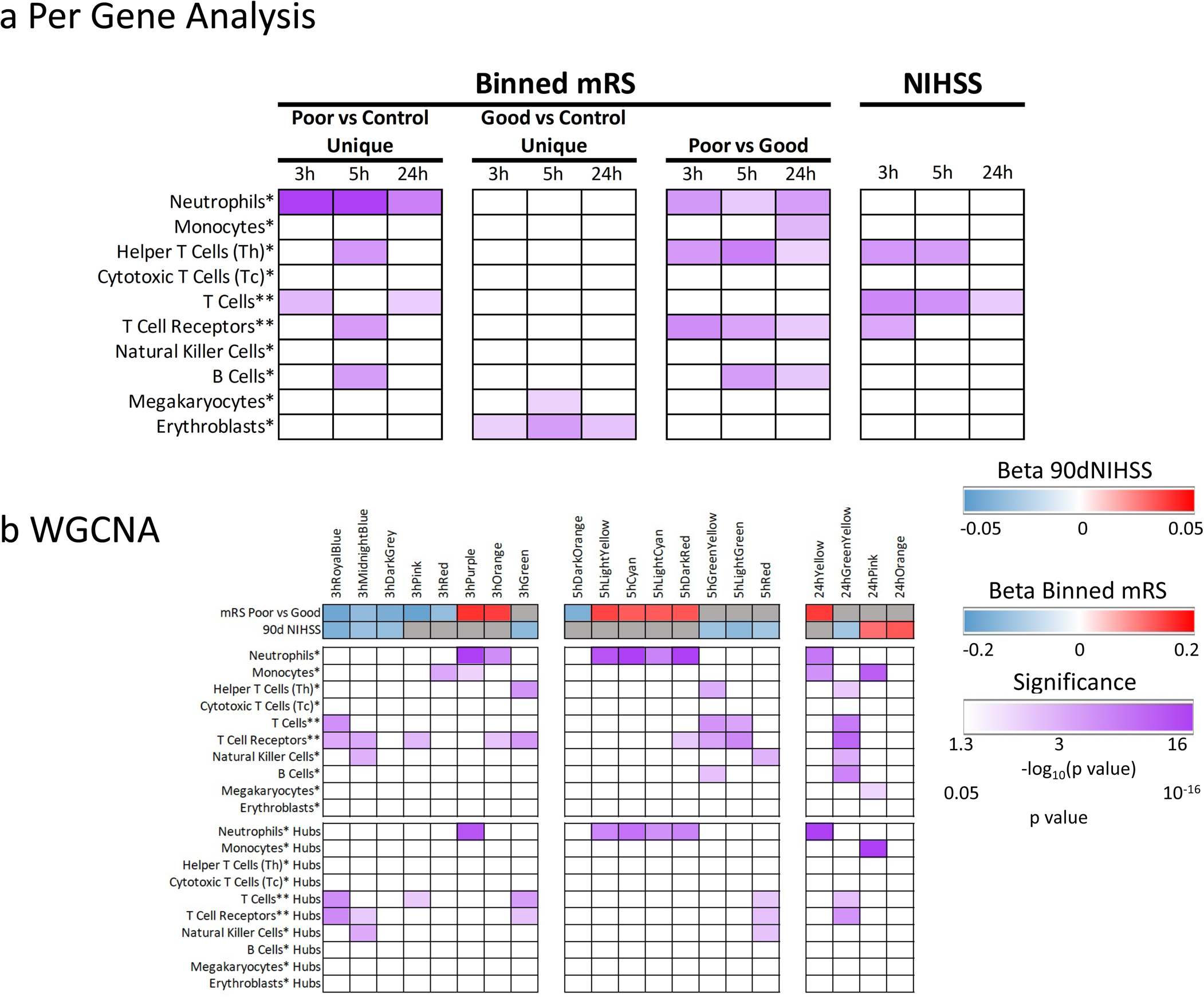
Enrichment in cell-type specific gene lists for the per-gene lists (a) and WGCNA modules (b). Purple shading represents –log_10_(*P* value) where 1.3 corresponds to a *P* value of 0.05. A higher – log_10_(p value) corresponds to lower (more significant - darker shades) *P* value. Non-significant hypergeometric probabilities are displayed as white cells. In panel (a) the results are based on genes differentially expressed in poor 90d mRS IS outcome vs. VRF controls, good 90d mRS IS outcome vs. VRF control, and genes correlating with 90d NIHSS. In panel (b) modules significant to 90d outcome (mRS poor vs. good, and NIHSS) are presented for the ≤3h Network, 5h Network, and 24h Network. Blue indicates down-regulated and red up-regulated expression with worse outcomes via the beta coefficient for outcome in a linear regression on the module eigengene. Grey indicates modules not significantly associated with the outcome measure. Enrichment of hub gene lists in cell-type specific lists are presented at the bottom. The single asterisk * indicates cell type list from Watkins et al. [34] and the double asterisk ** indicates the cell type list was from Chtanova et al. [33]. Some of the identified Neutrophil genes might be expressed by other granulocytes, i.e., basophils and eosinophils.

#### 3h-Gene Expression Associated with Good 90-day Functional Outcomes (mRS)

Eighty probe sets (representing 49 genes) were differentially expressed at ≤ 3h post IS in subjects with good outcome compared to VRFC (FDR-corrected *P*<0.05, FC> |2|) (Figure 1a, Table S1A). Of these, 20 probe sets were up-regulated and 60 down-regulated (Figure 1a, Table S1A). The 80 probe sets were overrepresented in 12 pathways (Figure 2a, Table S2A) with significant enrichment in Erythroblast-specific genes (3/49 genes (6.1%), *P*(overlap)LJ=LJ0.03) (Figure 3a).

#### Differential Expression ≤ 3h Post IS for Poor vs. Good 90-day Functional Outcome (mRS)

The data from these analyses is described in Supplementary Results (Poor vs. Good – 3h) and in Figures 1b, 3a, 4, and Tables S1A, S2A.

#### ≤3h Gene Expression Correlated with 90-day Outcome (NIHSS)

Six hundred seventy-one probe sets (538 genes) at ≤3h after IS were associated with 90d NIHSS (*P*<0.005). 469 probe sets negatively correlated and 202 positively correlated with 90d NIHSS (Figure 1c, Table S1A). The 671 probe sets were over-represented in 34 pathways (Figure S1, Table S2A) with significant enrichment with T helper cell-specific, T cell and T cell receptor signaling-specific genes (5/538 genes (0.9%), *P*(overlap)LJ= 1E-04; 21/538 (3.9%), *P*(overlap) = 8E-07; and 13/538 (2.4%), *P*(overlap) = 2E-03, respectively) (Figure 3A). All T cell receptor genes except *OSBPL10* negatively correlated with 90d NIHSS.

### Association of Gene Expression at 5h after IS with 90-day Stroke Outcome (mRS)

#### 5h-Gene Expression Associated with Poor 90-day Functional Outcome (mRS)

Seven hundred fifteen probe sets (526 genes) were differentially expressed at 5h in subjects with poor outcome compared to VRFC (FDR-corrected *P*<0.05, FC> |2| (Figure 1a). Of these, 431 probe sets were upregulated and 284 down-regulated (Figure 1a, Table S1B). The 715 probe sets were overrepresented in 106 pathways (Table S2B). Activated pathways included iNOS, Toll-like Receptor, IL-1, -6 and -7, NF-kB and STAT3 signaling; and regulation of IL-2 Expression was suppressed (Figure 2b, Table S2). There was enrichment in neutrophil-specific genes (76/526 genes (14.5%), *P*(overlap)LJ< 1E-16); in T helper cell and T cell receptor signaling-specific genes (5/526 genes (1.0%), *P*(overlap)=1E-04 and 14/526 genes (2.7%), *P*(overlap)=5E-04, respectively); and in B cell-specific genes (15/526 genes (2.9%), *P*(overlap)=6E-04) (Figure 3a). Most neutrophil-specific genes were up-regulated (74/76), while T cell-(14/19) and B cell-specific genes (14/15) were down-regulated in poor outcome subjects.

#### 5h-Gene Expression Associated with Good 90-day Functional Outcomes (mRS)

One hundred forty-six probe sets (100 genes) were differentially expressed at 5h after IS between good outcome and VRFC subjects (FDR-corrected *P*<0.05, FC > |2| (Figure 1a). Of these, 29 probe sets were up-regulated and 117 down-regulated (Figure 1a, Table S1B). The 146 probe sets were overrepresented in 9 pathways (Figure 2b, Table S2B), such as NRF2-Oxidative Stress Response and RhoGDI Signaling with significant enrichment in Erythroblast-specific genes (7/100 genes (7.0%), *P*(overlap)=7E-04) and Megakaryocyte-specific genes (4/100 (4.0%), *P*(overlap)=4E-02) (Figure 3a).

#### Expression at 5h Post IS for Poor vs. Good 90-day Functional Outcome (mRS)

The data from these analyses is described in Supplementary Results (Poor vs. Good – 5h) and in Figures 1b, 3a, 4, and Tables S1B, S2B.

**Fig. 4.**
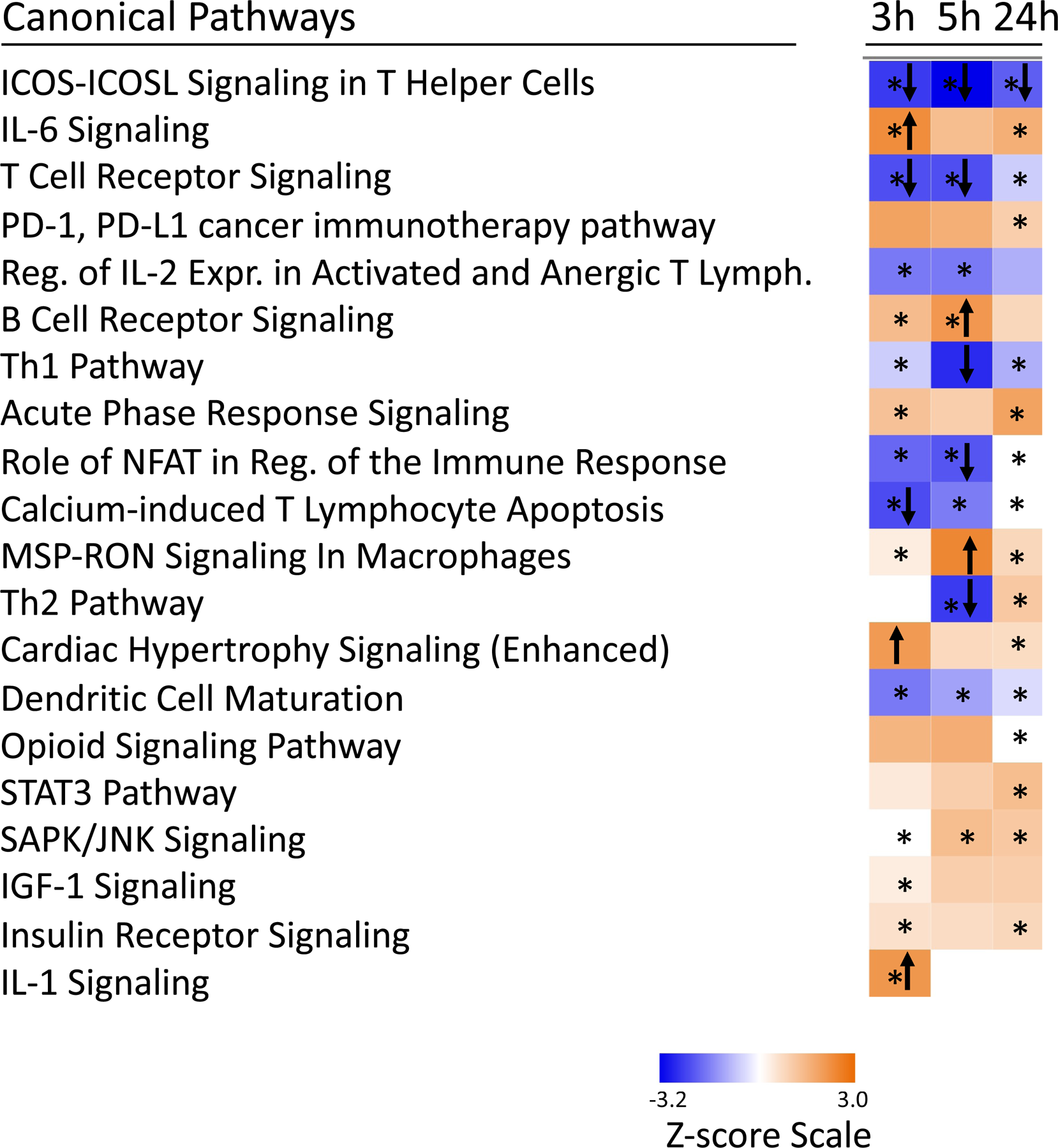
Pathway enrichment for Differentially Expressed Genes (DEGs) at ≤3h, 5h and 24h between subjects with poor 90-day mRS IS outcome compared to good 90d mRS IS outcome. The top 20 most significant activation or suppression relevant pathways are displayed. Blue bars indicate suppression / negative Z-score, and orange bars indicate activation / positive Z-score. Darker colors represent larger |Z-score|. ↑ (up arrow) represents Z ≥ 2, for the poor 90-day mRS IS outcome compared to good 90-day mRS IS outcome. ↓ (down arrow) represents Z ≤ -2 significant suppression in the poor 90-day mRS IS outcome compared to good 90-day mRS IS outcome. The asterisk * represents a statistically significant pathway (*P*<0.05). White cells represent non-significant functions and/or activity pattern prediction Z=0. Reg. – Regulation; Expr. – Expression; Lymph. – Lymphocytes.

#### 5h Gene Expression Correlated with 90-day Outcome (NIHSS)

Two hundred fifty-six probe sets (197 genes) at 5h after IS correlated with 90-day NIHSS (*P*<0.005). Of these 205 probe sets were negatively correlated and 51 were positively correlated (Figure 1c, Table S1B). The 256 probe sets were over-represented in 24 pathways with two suppressed pathways including Autophagy, and Regulation of IL-2 Expression in T Lymphocytes (Figure S1, Table S2B).

There was a significant enrichment in T helper-specific and T cell-specific genes (3/197 genes (1.5%), *P*(overlap)=8E-04 and 11/197 (5.6%), *P*(overlap)=2E-05), respectively (Figure 3a). T cell pathways negatively correlated with the 90-day NIHSS.

### Association of Gene Expression at 24h after IS with 90-day Stroke Outcome

The data for these analyses is provided in Supplementary Results (24h Correlation with 90-day mRS and NIHSS) and in Figures 1a, 1b, 1c, 2c, 3a, 4, S1 and Tables S1C-S2C.

### Weighted Gene Co-Expression Network Analysis Revealed Specific Gene Expression Modules Associated with Poor and Good 90-Day Outcomes Following IS

WGCNA was run on 28,686 Affymetrix probe sets for 36 IS subjects, with separate WGCNA runs generated for each time-point (≤3h, 5h, and 24h). Modules significantly associated with IS outcome such as 3hPurple, 3hRoyalBlue, 5hCyan, 24hYellow and 24hGreenYellow, and the canonical pathways significantly enriched in each module are presented in Figures 3b, 5a, 5b, S2, 6a, and 6b. Table 2 lists the hub genes for each of the modules and each of the time points.

**Fig. 5.**
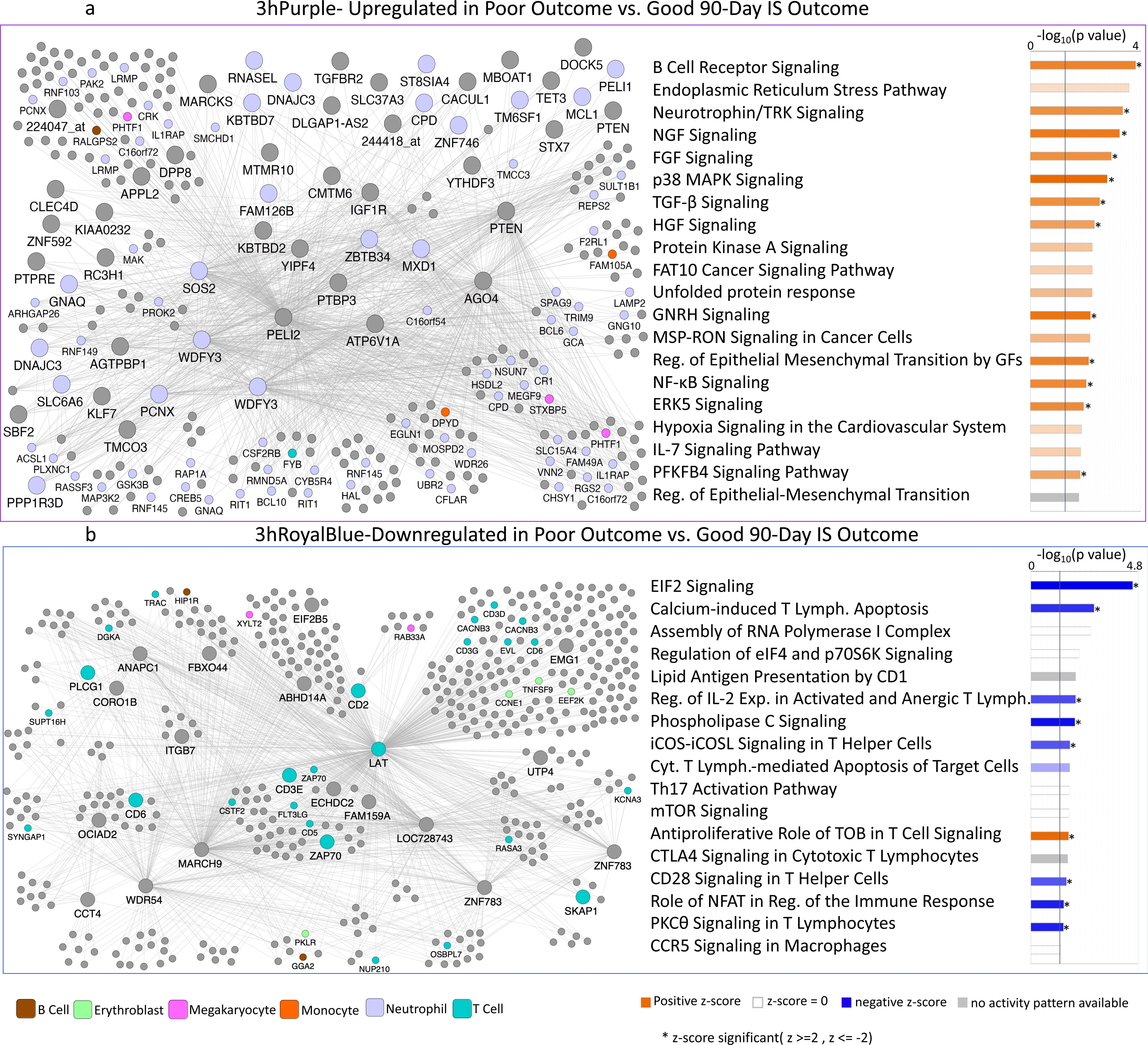
Network diagram (a left panel) and Pathway Enrichment (a right panel) for the outcome-significant (mRS poor vs. good) for the 3hPurple module. In the (a) left panel, the network diagram shows the connectivity of hubs and genes within the module. Nodes represent genes within the module and edges represent connections based on co-expression between genes. Larger nodes with large labels are hub genes, representing potential master regulators. Genes are grey by default and colored if they are cell type specific. In the (a) right panel, the top 20 most significant relevant pathways are displayed. The significance threshold (*P* = 0.05) corresponds to the vertical black line. Blue shading indicates suppression and orange activation with darker colors representing larger |Z-score|. Grey represents no activity pattern available for the pathway in the IPA knowledge base. An asterisk * represents statistically significant activation or suppression (Z ≥ 2 or Z ≤ -2) in poor outcome compared to good outcome. In (b) the Network diagram (b. left panel) and Pathway Enrichment (b. right panel) for the outcome-significant (mRS poor vs. good, 90-day NIHSS) for the 3hRoyalBlue module. The *LAT* gene is colored as T cell-specific though it is expressed in megakaryocytes and T Cells. White bars represent non-significant functions and/or activity pattern prediction Z=0. Other details of this figure are identical to those in (a). Reg. – Regulation; GFs. – Growth Factors; Expr. – Expression; Lymph. – Lymphocytes.

**Fig. 6.**
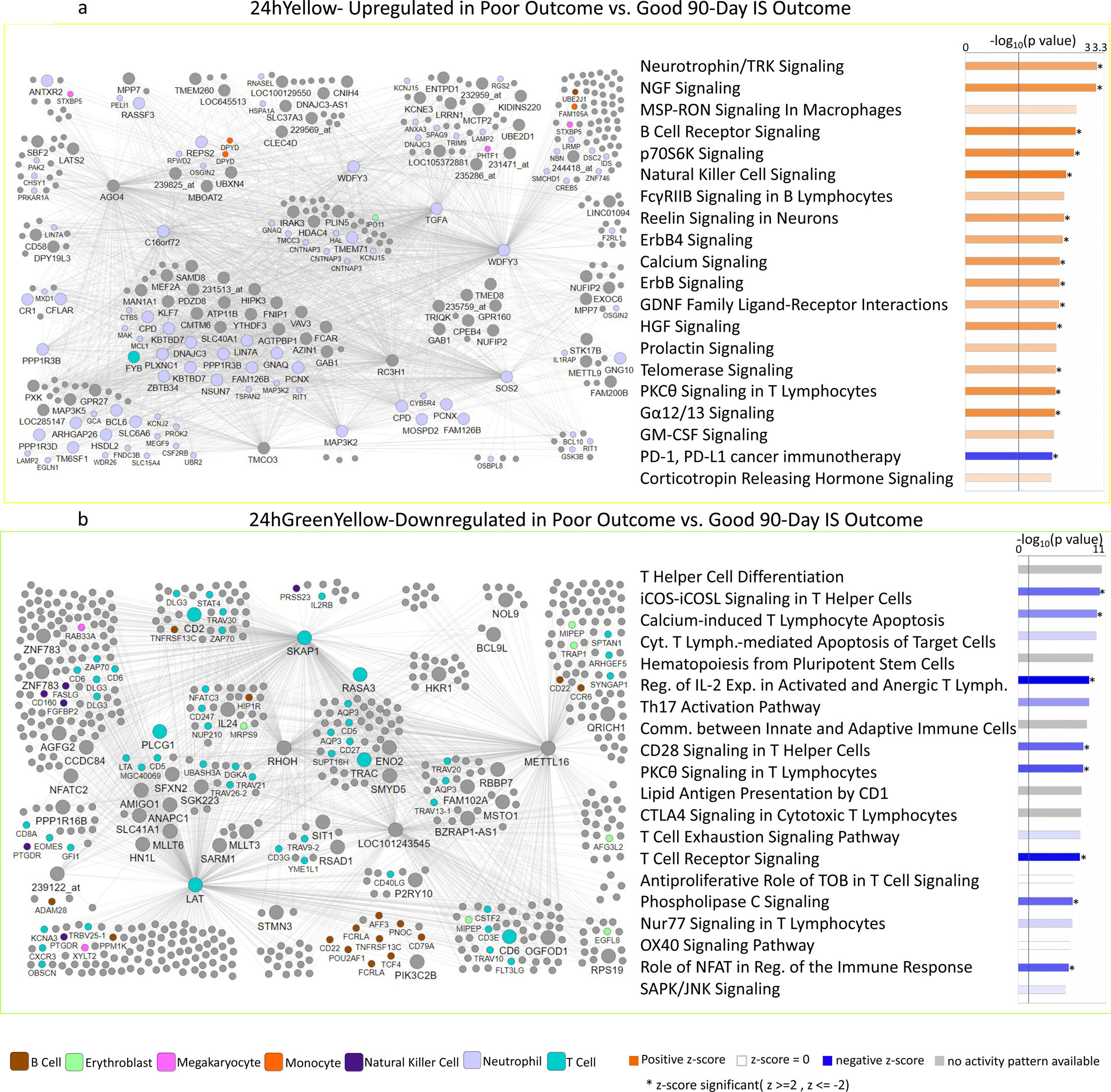
Network diagram (a left panel) and Pathway Enrichment (a right panel) for the outcome-significant (mRS poor vs. good) for the 24hYellow module. In the left panel, the network diagram shows the connectivity of hubs and genes within the module. Larger nodes with large labels are hub genes, representing potential master regulators. Genes are grey by default and colored if they are cell type specific. In the right panel, the top 20 most relevant significant pathways are displayed. The significance threshold (*P*= 0.05) corresponds to the vertical black line. Blue shading represents suppression and orange activation with darker colors representing larger |Z-score|. An asterisk * represents statistically significant activity pattern prediction with Z ≥ 2 or Z ≤ -2. In (b) the Network diagram (b left panel) and Pathway Enrichment (b right panel) for the outcome-significant (90-day NIHSS) for the 24hGreenYellow module. *IL2RB* and *CD247* are colored as T cell-specific but are also expressed in NK cells. *LAT* is colored as T cell specific, but also expressed in megakaryocytes. White bars represent non-significant functions and/or activity pattern prediction Z=0. Grey represents no activity pattern available for the pathway in the IPA knowledge base. Other aspects of this figure are identical to that described for panel (a). Cyt. – Cytotoxic; Reg. – Regulation; Expr. – Expression; Lymph. – Lymphocytes; Comm. – Communication.

**Table 2.**
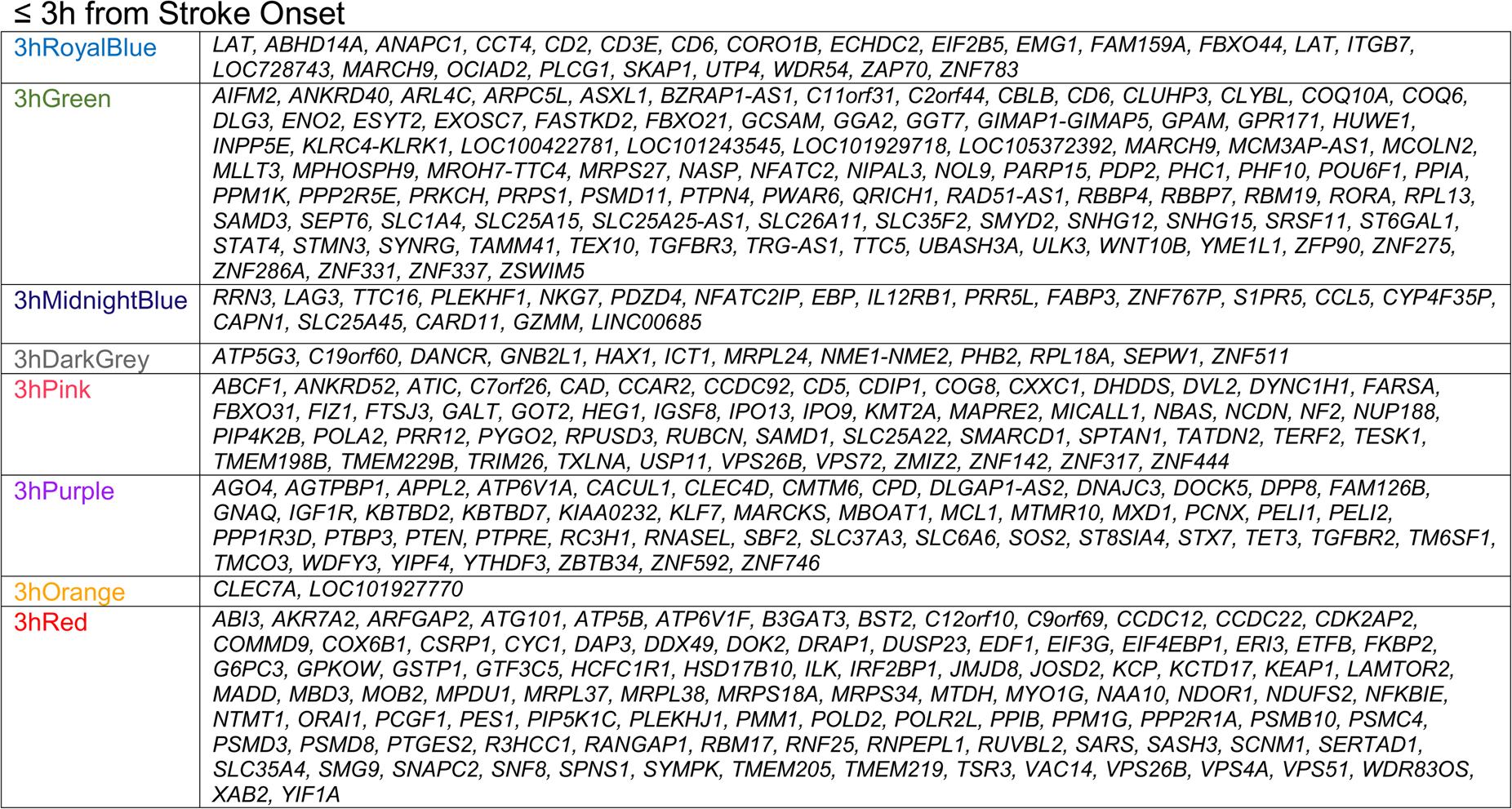
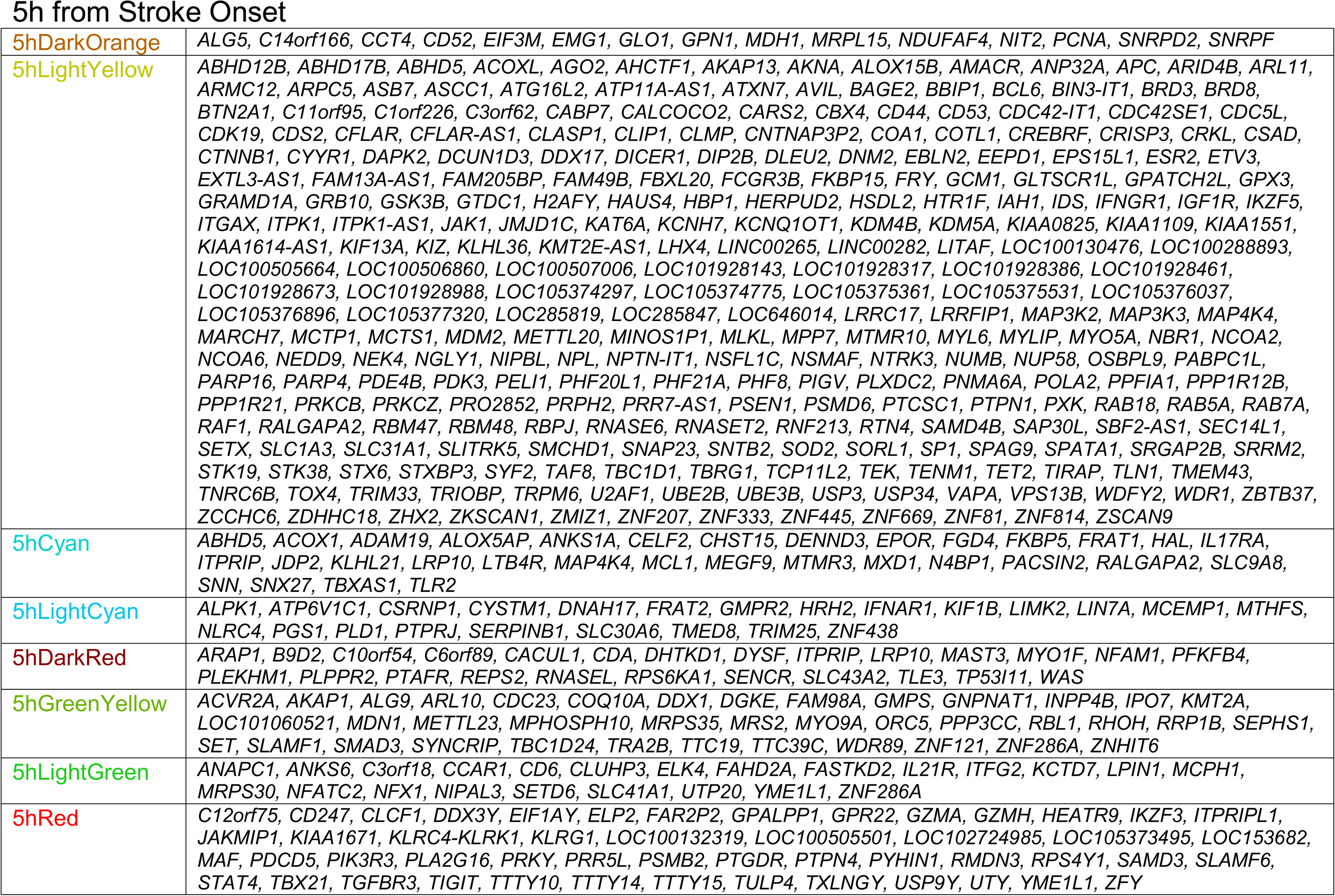
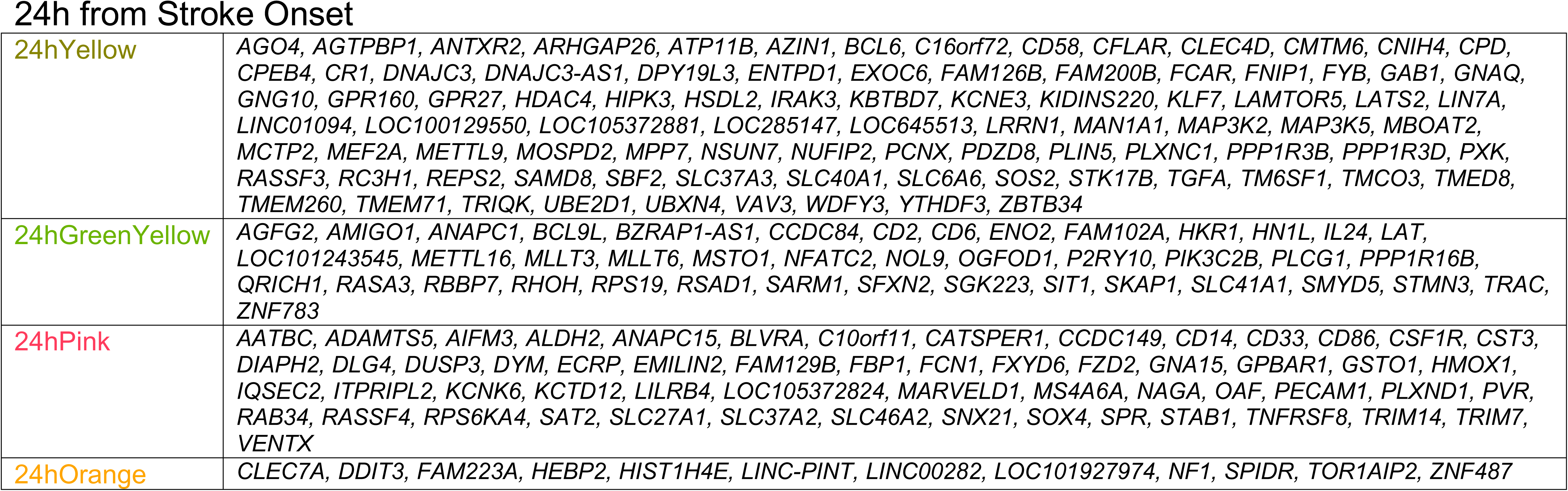
Hub genes in the outcome-significant modules. The probe sets without annotated genes are excluded.

#### Co-Expressed Gene Modules at ≤3h after IS Associated with 90-Day Outcome

Twenty-eight co-expressed probe set modules were identified for the ≤ 3h network (data not shown). Eight modules associated with 90-day Good and Poor outcomes (mRS) and/or 90-day NIHSS (Figure 3b). Six negatively correlated with 90-day outcome, including 3hRoyalBlue (Figure 3b and Figure 5b), 3hMidnightBlue and 3hDarkGrey which were significant for 90d mRS and 90d NIHSS; 3hPink for 90d mRS; and 3hGreen for 90d NIHSS (Figure 3b). The other two outcome-significant modules, 3hPurple (Figure 3b, Figure 5a) and 3hOrange, positively correlated with 90d mRS. Pathway analyses for each outcome-significant module are presented in Tables S4A. Most outcome-significant modules and/or their hubs (Table 2) were enriched in Neutrophil-, Monocyte-, T cell-, and/or NK cell-specific genes (Figure 3b). Neutrophil genes were enriched in positive-beta modules and/or hubs, while most T cell-genes and/or their hubs were enriched in negative-beta modules (Figure 3b).

The pathways over-represented in the T-cell receptor and other T-cell-specific hub genes at≤ 3h included T-cell pathways, calcium-induced T lymphocyte apoptosis, VEGF, NGF, neurotrophin/TRK and GDNF Signaling (Table S5A). Neutrophil-specific hub genes (3hPurple module) were enriched in 39 significant pathways such as IL-2, -6 and -7, and JAK/STAT Signaling (Table S5A). About half of the pathways (19/39) over-represented in neutrophil-specific hubs were also over-represented in the 108 pathways for T-cell specific hubs. However, the T-cell- and Neutrophil-specific hubs correlated in opposite directions with 90-day outcome (Figure 3b).

Figure 5a shows a 3h module enriched with neutrophil-specific genes (3hPurple) and Figure 5b for a 3h module enriched with T-cell specific genes (3hRoyalBlue) and their top over-represented pathways (right side of the figure). The 3hPurple module (TableS4A) for good vs. poor 90d mRS outcomes showed suppression of the PPARα/RXRα pathway which regulates NF-kB signaling. For the 3hRoyalBlue module T Cell Receptor pathways, IL-2 Regulation in T Lymphocytes, PKCθ Signaling in T Lymphocytes, and NFAT Regulation of Immune Responses were suppressed in Poor vs. Good outcome (Figure 5b, Table S4A).

#### Co-Expressed Gene Modules at 5h After IS Associated with 90-day Outcome

Thirty-two modules of co-expressed probe sets were identified within the 5h network (data not shown). Eight modules were associated with 90d Binned mRS and/or NIHSS (Figure 3b). Significant modules and/or their hubs were enriched in Neutrophil-, T cell-, NK cell-, and/or B cell-specific genes (Figure 3b). Notably, the four modules and their hubs enriched in neutrophil-specific genes (5hLightYellow, 5hCyan, 5hLightCyan, and 5hDarkRed) were positively associated with Poor vs. Good 90-day outcome (Figure 3b). The 5hCyan module, enriched in Neutrophil cell-specific genes and hub genes (Figure S2), was associated with 97 pathways (Figure S2 Right panel – top 20 relevant pathways, Table S4B). Among the top pathways were iNOS, B Cell Receptor, IL-7, ERK5 and FLT3 Signaling (Figure S2 Right panel). Also, the 5hLightGreen and 5hGreenYellow modules were enriched in T cell-specific genes that negatively correlated with 90d NIHSS had many associated pathways (Figure 3b, Table S4B). Pathway analysis of T cell specific hub genes showed 23 significant pathways including Th1, Th2, T Helper Cell Differentiation, and T Cell Exhaustion Signaling (Table S5B). Hub genes from modules positively correlated with 90-d mRS outcomes were enriched in neutrophil-specific genes (Figure 3b) with pathways including PI3K/AKT, JAK1, JAK2 and TYK2 in Interferon Signaling, STAT3, apoptosis and chemokine signaling (Table S5B). Inflammatory response disease functions including leukocyte migration at 5h were activated in Poor vs. Good outcome (data not shown). Ten hub genes including *ALOX5AP*, *DYSF*, *IFNAR1*, *IL17RA*, *MCL1*, *MYO1F*, *PELI1*, *PTAFR*, *RAF1*, and *RNASEL* were involved in the inflammatory response.

#### Co-Expressed Gene Modules at 24h After IS Associated with 90-day Outcome

Twenty-nine modules of co-expressed probe sets were identified in the 24h network (data not shown). Four were associated with 90d Binned mRS and/or NIHSS (Figure 3b). The significant modules and/or their hubs were enriched in Neutrophil-, Monocyte-, T cell-, NK cell and/or Megakaryocyte-specific genes (Figure 3b) and pathways (Table S4C). T cell-specific hub genes had 99 significant pathways including T Cell, Fc Epsilon RI, GP6, and ErbB4 signaling (Table S5C). The 24hYellow module was positively associated with 90-day mRS and was significantly enriched in neutrophil-specific genes (Figure 3b) and pathways (Table S4C). Pathway analysis of the Neutrophils-specific hub genes showed 76 significant pathways (Table S5C). The genes from 24hPink, 24hYellow and Hubs from 24hYellow were enriched in monocyte-specific genes. Monocyte specific hub genes showed 31 significant pathways including LPS/IL-1 Mediated Inhibition of RXR Function, IL-10, Macropinocytosis, HIF1α and mTOR signaling (Table S5C). The 24hYellow module is shown in Figure 6a along with 20 of its 88 significant pathways (Table S4C). Among the top relevant pathways, STAT3, Th1, FGF, ErbB4 and NF-κB Signaling were activated. The 24hGreenYellow module is shown in Figure 6b along with the top 20 of its 53 significant pathways (Table S4C).

### Predicting 90d Outcome from Changes in Gene Expression between 3h and 24h after IS

Since changes in gene expression over time may be more predictive of long-term outcome than gene expression at a single time point, we calculated the difference in gene expression between 24h and 3h (Δ(24h-3h)) post-IS. We derived ten genes (Table S6) that predicted 100% of good (n=18/18) and poor outcomes (n=7/7) in the training set. They also predicted 8/8 good 90-day mRS outcomes and 2/3 poor outcomes in the validation set (n=11; ROC-AUC= 0.88) (Figure 7). Thus, the 10 genes predicted 26/26 good outcomes and 9/10 poor outcomes overall.

**Fig. 7.**
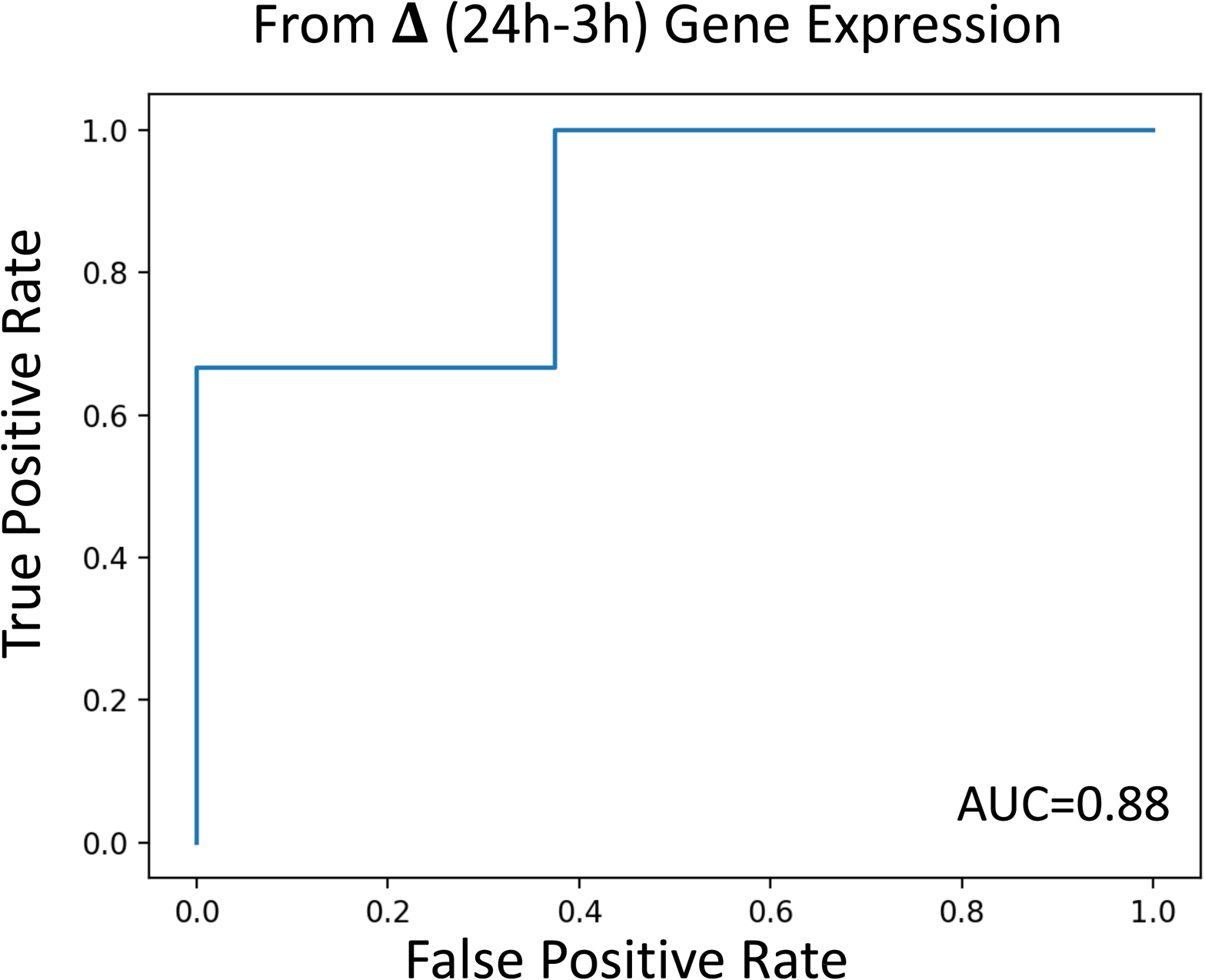
ROC Curve for Prediction of 90-day mRS outcome following Ischemic Stroke (IS). X-axis indicates the false positive rate, and the Y-axis indicates the True positive rate on the validation set. We used the difference in gene expression between 24 hours and 3 hours after IS in a logistic regression model to predict 90-day IS outcome. 10 genes predicted all subjects with good 90-day mRS outcome (8/8) and 2 out of the 3 subjects with poor outcome in the validation set (ROC-AUC= 0.88).

## Discussion

Expression of genes and gene co-expression modules in peripheral blood at early times after IS correlates with 90d outcomes. Upregulation of genes in neutrophils and down-regulation in monocytes, T cells and B cells may play a role in mediating damage and repair following stroke and ultimately affect long-term outcomes [37–39]. The findings expand our understanding of the transcriptomic changes in immune and clotting systems associated with outcome following human ischemic stroke. The identified genes may be novel targets for modulating outcome, and a subset of genes may predict outcome.

### Early Up-Regulated Immune/Inflammatory Genes/Pathways and Neutrophil-Specific Genes Associate with Poor 90-day Outcome

Inflammation plays a critical role in damage and repair following stroke [40]. Specific inflammatory blood markers correlate with outcomes after stroke [41], including pro-inflammatory cytokines like IL-1, IL-6, TNF, as well as anti-inflammatory cytokines like TGF and IL-10 [40–42]. Increases in matrix metalloproteinases (MMPs) including MMP-9 derived mainly from neutrophils are reported to cause BBB (blood-brain-barrier) damage [43, 44] and hemorrhagic complications [45–48]. MMP-9 levels correlate with infarct volume, stroke severity, and functional outcomes [46, 48]. In our study, *MMP9* expression (≤3h) was up-regulated 2.4 fold in subjects with poor 90d outcomes, which is consistent with other studies showing blood MMP-9 levels correlate with poor 90d IS outcomes [49, 50].

S100A12 mRNA, which is highly expressed by neutrophils, was up-regulated in this study at 3h in peripheral blood of subjects with poor 90d IS outcome (FC=2.1). Elevated S100A12 plasma levels at admission following IS have previously been associated with poor mRS outcome at 90-days [51]. In addition, S100A12 serum levels increase after traumatic brain injury (TBI) and intracerebral hemorrhage (ICH) [52, 53].

The STAT3 pathway was up-regulated at all three time-points, and several interleukin (IL)-related pathways (including IL-6 at 3h and 5h) were activated in subjects with poor IS outcomes. STAT3 promotes inflammatory responses [54] and IL-6 promotes phosphorylation of JAK2/STAT3 [55]. Serum IL-6 levels have previously been associated with poor long term IS outcomes [56, 57]. p38 MAPK, also significantly activated in subjects with poor 90-day IS outcome, modulates proinflammatory cytokines (IL-1β, TNF-α and IL-6) [58] and has been proposed as a therapeutic IS target [59].

*SMAD4* was up-regulated in IS subjects with poor 90d outcome at all three time points. SMAD4 has been implicated in inflammation and hypercoagulation in ischemic stroke [60], has been associated with BBB disruption [61] and in our previous study was up-regulated in IS subjects who later developed hemorrhagic transformation [60]. We have previously observed higher expression of *SMAD4* after IS, and particularly higher in individuals with the GG allele of rs975903 [62]. This could relate in part to post-translational regulation of SMAD proteins in response to TGF-β signaling [63].

Specific cytokine/chemokines such as *SPRED2*, *OSM* and *IL1A* (at all three time points), and *CXCL6* (at 3h post IS) were up-regulated in subjects with poor outcome, while *FLT3LG* and *CCR7* (at all three time points) were down-regulated. SPRED proteins modulate angiogenesis, vascular repair [64] and autophagy [65]. Thrombin aggravates astrocyte injury following IS by SPRED2 activation of autophagy pathways [65]. OSM (oncostatin M), an IL-6 cytokine family member, modulates inflammatory responses [66] and experimental stroke outcomes [66–68]. CXCL6 (C-X-C motif Chemokine Ligand 6), a chemoattractant for neutrophils and other granulocytes, is elevated following experimental ischemia-reperfusion injury [69]. *FLT3LG* (aka FLT-3L), down-regulated in 90d poor outcomes in our study, promotes differentiation of multiple hematopoietic cell lineages [70]. Low FLT3LF serum levels within 72 hours of stroke onset has been observed in severe stroke [70]. *CCR7* (C-C motif Chemokine Receptor 7), also down-regulated in poor outcomes, activates B and T lymphocytes and regulates T cell migration to sites of inflammation and stimulates dendritic cell maturation [71, 72].

Space precludes discussing most of the neutrophil genes also identified in the comparisons of poor vs. good outcomes, the WGCNA modules, and the hub genes associated with neutrophils. Overall, our findings in peripheral blood support the notion of complex effects of cytokines/chemokines on stroke pathophysiology and outcome. The early IS transcriptomic response in peripheral blood suggested a strong neutrophil response and activated inflammatory pathways associated with poor long-term outcome. However, inflammation has been shown to have detrimental as well as beneficial roles following IS that are highly time-dependent [73].

### Down-Regulation of Lymphocyte-Specific Genes Associated with Poor 90-day Outcome

Lymphocytes modulate ischemic stroke in a time-dependent manner [73], and the peripheral transcriptome responses of lymphocytes to ischemic stroke is fairly unique compared to other conditions [74]. In this study, we show enrichment in lymphocyte-specific genes for T cells, B cells and Natural Killer (NK) cells in the per-gene and WGCNA analyses. Most of these genes were down-regulated in poor 90-day outcome subjects.

Several over-represented T cell pathways were found in the outcome-significant WGCNA modules. Those included PKCθ Signaling in T Lymphocytes, CD28 Signaling in T Helper Cells, iCOS-iCOSL Signaling in T Helper Cells, Calcium-Induced T Lymphocyte Apoptosis, and the Th1 Pathway. T cell-specific hub genes in the modules included Cluster of Differentiation (*CD2*, *CD3E*, *CD5*, and *CD6*), *LAT*, *STAT3*, *STAT4*, *ZAP70*, *GZMM*, *PLEKHF1*, *PRKCH*, *TGFBR3*, *YME1L1*, *SPTAN1*, *DOK2*, *UBASH3A*, and *SKAP1*. Several of these genes have been implicated in stroke. For example, SKAP1 (Src kinase associated phosphoprotein 1) interacts with Src Family Kinases (SFKs) and stimulates T cell antigen receptors to activate integrins [75, 76]. The T cell-specific hub genes from the 3h WGCNA modules were enriched in pathways such as T Cell-Receptor Signaling and were predicted to be suppressed in subjects with poor 90-day outcomes. The T cell-specific hub genes included genes associated with stroke and ones important for repair after stroke (e.g., *AQP3*, *CD40LG*, *CD28*, *CAMK4*, *DNMT3A*, *EVL*, *KCNA3*, *LCK*, *MAL*, *PDE4D*, *SPTAN1*, *ARHGEF7*, *CBL*, *PLCG1*, *PRKCB*, *STAT1*, *STAT3*, *STAT5B*) [77]. *LCK* whose expression at 3h significantly associated with poor 90d mRS and NIHSS, is a member of SFK gene family expressed in T cells [33] and modulates outcomes in experimental ischemic stroke [78]. The cytokine *CD40LG* (CD40 ligand), also down-regulated in poor outcomes, is a target of the FDA-approved drug Letolizumab (PubChem BMS-986004). Our results could indicate that up-regulation of T cells and their genes might modulate long-term outcomes.

A brief discussion of the B cell and NK cell results is provided in the Supplementary Discussion (B cells and NK cells). Our data imply that down-regulation of lymphocyte-specific genes was associated within poor 90-day outcome. However, this is complicated by the fact that various studies have shown decreases of lymphocytes in blood of humans following stroke, thus partly accounting for decreases in expression. Moreover, since we investigated the changes in the transcriptome in whole blood, we can only infer cell-type specificity from known cell-specific gene expression and cannot decipher the entire transcriptomes of the specific lymphocyte cell types. Thus, additional studies of isolated peripheral blood cell types are needed to further refine the contribution of each cell type to the post-stroke response and its association with outcome.

### Coagulation, Platelet, and Cardiovascular Pathways Associated with Outcome IS

Coagulation and platelet activation are involved in causing IS and may play a role in long-term outcomes [79, 80]. In our study poor 90d outcomes were associated with enrichment of Thrombin pathways at 5h and 24h, Thrombopoietin Signaling at 5h and the Intrinsic Prothrombin Activation Pathway at 24h after stroke. Thrombopoietin (TPO), protective in experimental focal stroke [81], stimulates the production and differentiation of megakaryocytes and regulates platelet formation. In addition, several coagulation factors such as Factor 5 (*F5*, coagulation factor V; causative gene in Factor V Leiden thrombophilia), *F8* and *F12*, which are part of the Intrinsic Prothrombin Activation Pathway, were up-regulated at 24h in IS subjects with poor 90d outcomes. These findings support suggestions coagulation and fibrinolysis biomarkers are predictive of thrombolysis treatment outcome after IS [82]. Though our results provide evidence that early activation of peripheral coagulation pathways associate with long-term outcome, it is unclear whether this relates only to early fibrinolysis or to other effects on brain repair during recovery. Thus, further studies into their potential usefulness as treatment targets are warranted.

Cardiovascular function pathways activated at 5h post stroke that correlated with 90d outcomes included Adrenomedullin signaling pathway, Renin-Angiotensin Signaling and HIF1α Signaling. Plasma Adrenomedullin levels increase following IS and are an independent predictor of 3-month IS outcomes [83, 84]. HIF1α Signaling, also activated at 5h in poor mRS outcome, regulates most hypoxia responsive genes [85]. HIF1α serum levels correlate with worse IS outcomes [86]. Renin-Angiotensin Signaling, modulated at 3h, 5h and 24h, was associated with 90d poor outcome subjects. Renin-angiotensin (RAS) contributes to increased arterial pressure and has been associated with local cerebrovascular dysfunction [87]. In ischemic stroke there may be an imbalance in the two opposing axes of RAS – a ‘classical axis’ and ‘alternative axis’ mediated by Angiotensin II and Angiotensin-(1–7), respectively [88]. Modulating RAS influences experimental stroke outcomes [89]. Thus, our human data suggest early changes of coagulation and cardiovascular function pathways are associated with poor long-term outcomes, and thus may be therapeutic targets.

### Growth Factor Signaling

Several growth factor signaling pathways were enriched in subjects with poor mRS outcomes, including Erythropoietin, Fibroblast Growth Factor (FGF), Transforming growth factor-β (TGF-β), Growth Hormone, Granulocyte-Macrophage Colony-Stimulating Factor (GM-CSF), Hepatic Growth Factor (HGF), VEGF and VEGF Family Ligand-Receptor signaling. Several studies have found levels of certain growth factors correlate with good outcomes after IS [90–92]. However, growth factors can have pleiotropic and sometimes opposing effects [93, 94]. For example, higher VEGF levels exacerbated hemorrhage after experimental brain arteriovenous malformations [94] and can worsen edema in experimental IS [95]. Another study showed high blood levels of FGF23 increased the risk for cardiovascular disease and stroke [93]. We previously found several growth factor signaling pathways associated with larger ICH volumes and peri-hematomal edema volumes [28]. Further studies are needed to better understand the association between early changes in growth factor signaling and long-term outcome.

### Module Hubs

Hubs, the most inter-connected genes in each co-expression module defined in WGCNA, are potential master regulators of gene expression. Five outcome-significant modules are highlighted in Figures 5, 6 and S2: two modules being enriched with T-cell specific genes and three with neutrophil-specific genes.

Hub genes in neutrophil modules included *AGO4* (Argonaute RISC Component 4) and *PTEN* (Phosphatase And Tensin Homolog). These have been implicated in immune-related pathways and brain injury [96–98]. The Argonaute RISC Component family responds to hypoxia [96] which can contribute to poor functional outcome [97]. PTEN protects against cerebral ischemia [98]. Another neutrophil hub gene, *PELI2*, which contributes to microglial activation following subarachnoid hemorrhage [99], could have a similar role in IS. *MEGF9,* a neutrophil-specific hub gene, could also impact IS outcome [100]. *PPP1R3B*, also a neutrophil-specific hub gene, has polymorphisms associated with serum LDL-C levels that contribute to coronary artery and IS disease risk [101].

T cell-specific hub genes included *ZAP70*, *LAT*, *SKAP1*, *PLCG1*, and *CD3E*. LAT protein is phosphorylated by ZAP70/Syk protein tyrosine kinases following activation of the T-cell antigen receptor (TCR) transduction pathway [102]. *ZAP70* is differentially expressed at 3h and 24h in poor outcome subjects, and is up-regulated in ICH patients [103]. *PLCG2* is expressed in human and mouse brain microglia. PLC enzymes like PLCG1 are key elements in signal transduction networks, with the PLCG2 P522R variant being protective against AD [104]. *SKAP1* encodes a T cell adapter protein that promotes adhesion and degranulation, which stimulates T cell antigen receptors to activate integrins. Given the large number of hub genes identified across multiple cell types, however, there is a need to develop approaches for determining which might be the best treatment targets.

### Predicting 90-day Outcome from Gene Expression Following IS

Though a previous study showed that gene expression surrogates could predict improvement in NIHSS from admission to discharge [5], this is the first study to use early gene expression to predict 90d mRS and NIHSS. We engineered a new feature of change of expression between 24h and 3h to predict 90d outcomes. We identified 10 genes which predicted 18/18 good outcome and 7/7 poor 90d outcomes in a training set, and 8/8 good outcome and 2/3 poor 90d mRS outcomes in a validation set. Among the 10 predictors were *AVPR1A* (arginine vasopressin receptor 1A), a receptor for arginine vasopressin (AVP), which mediates platelet aggregation and release of coagulation factors, exacerbates brain inflammatory responses to injury and promotes BBB disruption and increases cerebral edema in brain injury. AVPR1A increases in injured brain, plasma and cerebrospinal fluid in IS, ICH, subarachnoid hemorrhage and TBI patients [105]. A SNP in another gene in the 10-gene predictor set, *MSRB3*, is associated with increased odds of stroke in Alzheimer’s Disease [106]. Another predictor was *APCDD1*, a Wnt/β-catenin Signaling inhibitor, which coordinates vascular remodeling and barrier maturation of retina blood vessels [107].

Another gene was *HIPK2*, which is a serine/threonine-protein kinase involved in the hypoxia response as a transcriptional co-suppressor of *HIF1A* [108]. Silencing the circular RNA form of HIPK2 in neural stem cells improved functional recovery post IS [108]. The top over-represented pathway in the 10-classifier gene set was p53 Signaling which is implicated in the regulation of cell death in stroke [109, 110].

Our approach of calculating the change in gene expression between 3h and 24h post IS improved sensitivity compared to 24h alone (data not shown). Though the accuracy for predicting good 90-day outcome is excellent, the accuracy for predicting poor outcome in the validation set was modest due to very small sample size. Nevertheless, in this study the classifier consisting of gene expression data alone had a significantly higher accuracy than a classifier where age, sex and 24h NIHSS [111] were used as predictors (data not shown). A model consisting of gene expression plus age, sex and 24h NIHSS did not improve the 90d outcome prediction in comparison to using gene expression alone (data not shown). The results demonstrate feasibility of developing gene predictors of IS outcome, though the best predictors may differ somewhat once large sample sizes are analyzed.

### Limitations

The findings from this study need to be validated in larger cohorts. The cell-specific genes used here [33, 34] were identified in healthy subjects and may change expression pattern with disease. In addition, since changes in cell count of specific peripheral blood cell types have been reported following ischemic stroke, some of the outcome differences of expression in this study could be due to changes in different proportions of cells.

The study included repeat blood draws of ischemic stroke subjects at pre-treatment (3h) and post-treatment (5h, 24h) time points. Since no IS subjects were untreated, the results cannot tease out the contribution of treatment to outcome. Nevertheless, the results provide interesting insights. First, genes at times pre- and post-treatment correlated with 90d outcomes, with the greatest number of genes identified pretreatment (3h). Second, comparison of the outcome genes to our previous study of tPA responsive genes in blood of rats with strokes from Jickling *et al*. [112] shows very little overlap except for 5h good outcome vs. VRFC genes, suggesting most of 5h and 24h outcome genes were not related to tPA administration (Overlap genes in Table S7). Third, the best predictor of 90d IS outcome was the change of gene expression from the pre-treatment time point (3h) to the 24h post-treatment time point, suggesting gene expression both before and after treatment have predictive value. Indeed, future clinical trials might use the model here where gene expression in blood both prior to and after treatment could be used to predict 90d outcomes.

## Data Availability

The data from this study are available upon reasonable request.

## Acknowledgements

This study was funded by the NIH (National Institutes of Health) NINDS (National Institute of Neurological Disorders and Stroke) R01 (NS106950, NS075035, NS097000 and NS101718 -FRS, BSS, BPA), and by the American Heart Association (16BGIA27250263-BSS). GCJ receives research funding from CIHR, Heart and Stroke Foundation of Canada, University Hospital Foundation and CRC.

## Compliance with Ethical Standards and Conflicts of Interest

The authors report no competing financial interests and declare no conflicts of interest. The CLEAR clinical trial was IRB approved by the University of Cincinnati, and informed consent from the patient or their proxy was obtained on every subject. The SAVVY trial was approved by the Wake Forest University IRB and informed consent was obtained from every patient or their proxy.

## Data Availability

The data from this study are available upon reasonable request.

## Author Contributions

The study and analyses were designed by HA, FRS and BSS. Data analyses and review were performed by HA and BSS. The manuscript was written by HA, FRS and BSS. All authors reviewed the study and manuscript, provided input on its contents, and agreed to its content.

## Supplemental Figures

**Fig.S1** Pathway enrichment presented for genes whose expression correlates with 90-day NIHSS at ≤3h, 5h and 24h. The top 20 most significant activation or suppression relevant pathways are displayed. Blue shading indicates suppression (negative Z-score), orange indicates activation (positive Z-score), and darker colors represent larger |Z-score|. ↑ (up arrow) represents Z ≥ 2, significant activation and ↓ (down arrow) represents Z ≤ -2 significant suppression in subjects with worse outcome compared to subjects with better 90-day outcome. The asterisk * represents a statistically significant pathway (*P*<0.05 is significant). White cells represent non-significant functions and/or activity pattern prediction Z=0. Grey represents no activity pattern available for the pathway in the IPA knowledge base. Reg. – Regulation; Expr. – Expression; Lymph. – Lymphocytes; Cyt. – Cytotoxic.

**Fig.S2** Network diagram (left panel) and Pathway Enrichment (right panel) for the 5hCyan module which is significant for association with 90-day mRS. The left panel network diagram shows the connectivity of hubs and genes within the module. Larger nodes with large labels are hub genes, representing potential master regulators. Genes are grey by default and colored if they are cell type specific. In the right panel, the top 20 relevant significant pathways are displayed, with the vertical line indicating a *P*=0.05. Blue shading indicates suppression (negative Z-score) and orange indicates activation (positive Z-score), and darker color represents larger |Z-score|. The asterisk * represents Z ≥ 2 or Z ≤ -2 in poor outcome compared to good outcome. Signal. – Signaling.

### Supplementary Tables

**Table S1A.** Gene expression of genes **at** ≤**3 hours** of ischemic stroke (IS) onset that associates with 90-day outcome

• 467 Genes significantly differentially expressed between Poor 90-day mRS IS Outcome and Vascular Risk Factor (VRF) control (FDR <0.05, |FC|>2)
• 49 Genes significantly differentially expressed between Good 90-day mRS IS Outcome and VRF control (FDR <0.05, |FC|>2)
• 709 Genes (at ≤3 hours) significantly differentially expressed between subjects with Poor 90-day mRS IS Outcome and subjects with Good 90-day mRS Outcome (*P*<0.05, |FC|>1.3)
• 538 Genes significantly associated with 90-day NIHSS Outcome (*P*<0.005)

**Table S1B.** Gene expression of genes **at 5 hours** of IS onset that associates with 90-day outcome

• 526 Genes significantly differentially expressed between Poor 90-day mRS IS Outcome and VRF control (FDR <0.05, |FC|>2)
• 100 Genes significantly differentially expressed between Good 90-day mRS IS Outcome and VRF control (FDR <0.05, |r|>2)
• 658 Genes significantly differentially expressed between Poor 90-day mRS IS Outcome and Good 90-day mRS Outcome (*P*<0.05, |FC|>1.3)
• 197 Genes significantly associated with 90-day NIHSS Outcome (p <0.005)

**Table S1C.** Gene expression of genes **at 24 hours** of IS onset that associates with 90-day outcome

• 571 Genes significantly differentially expressed between Poor 90-day mRS IS Outcome and VRF control (FDR <0.05, |FC|>2)
• 35 Genes significantly differentially expressed between Good 90-day mRS IS Outcome and VRF control at 24 hours (FDR<0.05, |FC|>2)
• 363 Genes significantly differentially expressed between Poor 90-day mRS IS Outcome and Good 90-day mRS IS Outcome (*P*<0.05, |FC|>1.3)
• 147 Genes significantly associated with 90-day NIHSS Outcome (p<0.005)

**Table S2A.** IPA Canonical Pathway Enrichment (*P*<0.05) for gene expression **at** ≤**3 hours** of ischemic stroke (IS) onset that associates with 90-day outcome

• IPA Canonical Pathway Enrichment for 467 Genes Significant to Poor 90-day mRS Outcome as compared to VRF Controls
• IPA Canonical Pathway Enrichment for 49 Genes Significant to Good 90-day mRS Outcome as compared to VRF Controls
• IPA Canonical Pathway Enrichment for 709 Genes Significant to Poor 90-day mRS Outcome vs. Good 90-day mRS Outcome
• IPA Canonical Pathway Enrichment for 538 genes associated with 90-day NIHSS Outcome **Table S2B.** IPA Canonical Pathway Enrichment (*P*<0.05) for gene expression **at 5 hours** of IS onset that associates with 90-day outcome
• IPA Canonical Pathway Enrichment for 526 Genes Significant to Poor 90-day mRS Outcome as compared to VRF Controls
• IPA Canonical Pathway Enrichment for 100 Genes Significant to Good 90-day mRS Outcome as compared to VRF Controls
• IPA Canonical Pathway Enrichment for 658 Genes Significant to Poor 90-day mRS Outcome vs. Good 90-day mRS Outcome
• IPA Canonical Pathway Enrichment for 197 genes associated with 90-day NIHSS Outcome

**Table S2C.** IPA Canonical Pathway Enrichment (*P*<0.05) for gene expression **at 24 hours** of IS onset that associates with 90-day outcome

• IPA Canonical Pathway Enrichment for the 571 Genes Significant to Poor 90-day mRS Outcome as compared to VRF Controls
• IPA Canonical Pathway Enrichment for the 35 Genes Significant to Good 90-day mRS Outcome as compared to VRF Controls
• IPA Canonical Pathway Enrichment for the 363 Genes Significant to Poor 90-day mRS Outcome vs. Good 90-day mRS Outcome
• IPA Canonical Pathway Enrichment for the 147 genes associated with 90-day NIHSS Outcome

**Table S3A.** DAVID Gene Ontology Enrichment (FDR *P*<0.05) for the 467 Genes (at ≤3 hours) Significant to Poor 90-day Outcome as compared to VRF Controls

**Table S3B.** DAVID Gene Ontology Enrichment (FDR *P*<0.05) for the 571 Genes (at 24 hours) Significant to Poor 90-day Outcome as compared to VRF Controls

**Table S4A.** IPA Canonical Pathway Enrichment (*P*<0.05) for outcome-significant WGCNA modules using gene expression **at** ≤**3 hours** from IS onset

• 3hDarkGrey Significant to NIHSS at 90day
• 3hDarkGrey Significant to mRS at 90day
• 3hGreen Significant to NIHSS at 90day
• 3hMidnightBlue Significant to NIHSS at 90day
• 3hMidnightBlue Significant to mRS at 90day
• 3hOrange Significant to mRS at 90day
• 3hPink Significant to mRS at 90day
• 3hPurple Significant to mRS at 90day
• 3hRed Significant to mRS at 90day
• 3hRoyalBlue Significant to mRS at 90day
• 3hRoyalBlue Significant to NIHSS at 90day

**Table S4B.** IPA Canonical Pathway Enrichment (*P*<0.05) for outcome-significant WGCNA modules using gene expression **at 5 hours** from IS onset

• 5hCyan Significant to mRS at 90day
• 5hDarkOrange Significant to mRS at 90day
• 5hDarkRed Significant to mRS at 90day
• 5hGreenYellow Significant to NIHSS at 90day
• 5hLightCyan Significant to mRS at 90day
• 5hLightGreen Significant to NIHSS at 90day
• 5hLightYellow Significant to mRS at 90day
• 5hRed Significant to NIHSS at 90day

**Table S4C.** IPA Canonical Pathway Enrichment (*P*<0.05) for outcome-significant WGCNA modules using gene expression **at 24 hours** from IS onset

• 24hGreenYellow Significant to NIHSS at 90day
• 24hOrange Significant to NIHSS at 90day
• 24hPink Significant to NIHSS at 90day
• 24hYellow Significant to mRS at 90day

**Table S5A.** IPA Canonical Pathway Enrichment (*P*<0.05) for some cell specific WGCNA hubs at ≤ **3 hours**

• T cell receptor and other T cell-specific Hub Genes
• Neutrophil-specific Hub Genes

**Table S5B.** IPA Canonical Pathway Enrichment (*P*<0.05) for some cell specific WGCNA hubs at **5 hours**

• T cell receptor and other T cell-specific Hub Genes
• Neutrophil cell-specific Hub Genes

**Table S5C.** IPA Canonical Pathway Enrichment (*P*<0.05) for some cell specific WGCNA hubs at **24 hours**

• T cell receptor and other T cell-specific Hub Genes
• Neutrophil cell-specific Hub Genes
• Monocyte cell-specific Hub Genes

**Table S6.** The 10 genes used as predictors of Poor and Good 90-day mRS Outcomes

**Table S7.** Genes regulated by tPA in a rat stroke model (Jickling *et al*., 2010) overlapped with the 5h and 24h outcome genes in this study. The only significant overlap of tPA genes with outcomes genes was for 5h good outcome vs VRFC (*P*=0.005).

