## Supplemental Material for "Peripheral Blood Gene Expression at 3 to 24 Hours Correlates with and Predicts 90-Day Outcome Following Human Ischemic Stroke"

for

**Supplemental Methods**

Supplementary Methods WGCNA1. All networks were constructed using the same 36 CLEAR trial IS patients and 28,686 probe sets. Data was imported into R and checked for missing or zero-variance probe sets using the function *goodSamplesGenes*. Pearson correlations were used to measure co-expression [1]. An approximate scale-free topology was depicted by the data. Soft thresholding powers (β) of 6, 9, and 7 were selected for the ≤3 hours network, 5 hours network, and 24 hours network, respectively, to maximize strong correlations between genes and minimize weak correlations [2]. A signed network was used to consider both positive and negative correlations [3]. The *cutreeDynamic* function (method = tree, deepsplit = 1; minimum module size = 150) was used to form modules because it can identify nested modules in complex dendrograms [4]. Hub genes, the top 5% most interconnected genes in each module, were identified by their interconnectivity (kIN—the gene’s intramodular connectivity). Hub genes may be potential master regulators within their networks [5,6].

Supplementary Methods WGCNA2. Module association with Binned mRS (Poor 90-day Outcome, Good 90-day Outcome) was modeled by Y*_i_* = *μ* + Binned_mRS + Hypercholesterolemia + Hypertension + Diabetes + Group + Age + Sex + ε*_i_* where Y*_i_* is the module eigengene value (first principal component of expression), *μ* is the common effect for the whole experiment, Binned_mRS is a binary categorization of the 90-day patient outcome into Good (mRS = 0-2) and Poor (mRS =3-5), Group is the patient treatment group (tPA; tPA+eptifibatide). Module association with 90-day NIHSS as a continuous variable was modeled by Y*_i_* = *μ* + baseNIHSS + 24hNIHSS + 5dNIHSS + 90dNIHSS + Hypercholesterolemia + Hypertension + Diabetes + Group + Age + Sex + ε*_i_*. A value of *P* < 0.05 was considered significant.

Supplementary Methods Pathway Analyses. IPA’s pathway activity prediction analysis determined if the significant pathways were activated or inhibited using the expression direction (correlation coefficient or fold change) of the input genes. IPA’s Z-score algorithm calculated the predicted overall activation/inhibition states of the canonical pathways by statistically comparing our uploaded datasets with the IPA knowledge base’s expression patterns [7]. For the network analyses results, since we calculated the correlation or the fold change between the 90-day NIHSS outcome and Binned mRS (respectively) with the module’s eigengene, and since co-expressed probe sets within each module may have different direction correlations or fold changes, we separately calculated the partial correlation and fold change (contrast Poor 90-day Outcome vs. Good 90-day Outcome) between each probe set and the particular outcome measure using the same model used to assess module significance. This correlation or fold change value was input into IPA for prediction of the pathways’ activation/suppression status. Canonical pathways with Z ≥ 2 were considered significantly activated, while ones with Z ≤ −2 were considered significantly suppressed. For modules associated with Binned mRS, since we input fold change for Poor vs. Good outcome, pathways predicted to be activated are predicted to be activated in subjects who will have poor 90-day outcomes compared to those with good outcomes, while pathways predicted to be suppressed are predicted to be suppressed in subjects who will have poor outcomes compared to subjects with good outcomes. Similarly, for modules associated with 90-day NIHSS, since we input correlation coefficients with NIHSS, where the higher the NIHSS value, the worse the outcome, pathways with significant activation mean that the more activated the pathways, the worse the 90-day outcome. Pathways with significant suppression mean that the more suppressed the pathway, the worse the 90-day outcome.

IPA and the NIAID/NIH DAVID Bioinformatics Resources ([http://david.abcc.ncifcrf.gov](http://david.abcc.ncifcrf.gov/)) were used to identify statistically significant functional categories in the data set using a modified Fisher's exact test (*P* <0.05). The threshold for the EASE score used for the gene-enrichment analysis is based on a modified Fisher's exact *P* value. Fisher's exact tests determined whether there were more genes per biological category that are differentially expressed between the groups than would be expected by chance.

**Supplemental Results**

Supplementary Results (Poor vs. Good - 3h)

One thousand twenty-seven probe sets (representing 709 genes) were differentially expressed between subjects with poor and good 90-day mRS outcome with *P* <0.05 and a fold change (FC)> |1.3|. 432 probe sets were downregulated, 595 were upregulated in poor vs. good outcome patients (Figure 1b, Table S1A). The 1027 probe sets were over-represented in 62 pathways, with three activated (IL-1, IL-6 signaling and Remodeling of Epithelial Adherens Junctions) and three suppressed (ICOS-ICOSL Signaling in T Helper Cells, Calcium-induced T Lymphocyte Apoptosis, and T Cell Receptor Signaling) (Figure 4 and Table S2A). In addition, there was a significant enrichment in neutrophil-specific genes (46/709 genes (6.5%), *P*(overlap) = 2E-04); and in T helper cell-specific and T cell receptor and signaling-specific genes (5/709 genes (0.7%), *P*(overlap)=4E-04 and 21/709 genes (3.0%), *P*(overlap)=4E-06, respectively) (Figure 3a). Notably, 45/46 neutrophil-specific genes were up-regulated, while 20/26 T cell-specific genes were down-regulated in subjects with poor compared to subjects with good 90-day functional outcome.

Supplementary Results (Poor vs. Good – 5h)

Nine hundred thirty-one probe sets (representing 658 genes) were differentially expressed between poor outcome and good 90-day mRS outcome with *P* <0.05 and FC > |1.3| (Figure 1b). Of these, 508 were down-regulated, and 423 up-regulated in poor vs. good outcome (Figure 1b, Table S1B). They were over-represented in 56 pathways (Table S2B), with two activated including B Cell Receptor Signaling and five suppressed (ICOS-ICOSL Signaling in T Helper Cells, Th2 Pathway, and T Cell Receptor Signaling, and Role of NFAT in Regulation of the Immune Response). Several T cell-related pathways were overrepresented in the gene list (Table S2B, Figure 4). In addition, there was significant enrichment in neutrophil-specific genes (35/658 genes (5.3%), *P*(overlap)=0.02); B cell-specific genes (17/658 genes (2.6%), *P*(overlap) = 8E-04); and T helper-specific and T cell receptor and signaling-specific genes (8/658 genes (1.2%), *P*(overlap)=6E-08 and 15/658 genes (2.3%), *P*(overlap)=1E-03, respectively) (Figure 3a). The neutrophil-specific genes were up-regulated in poor outcome (except *CCR3*), while T cell-specific genes were down-regulated in poor outcome, except five genes-*SOS2*, *CBL*, *SNTB2*, *APBB1IP* and *PRKCB*, which were up-regulated in poor outcome.

**Supplementary Results (24h Correlation with 90-day Outcome (mRS and NIHSS)**

*24h Gene Expression Associated with Poor 90-day Functional Outcome (mRS)*

Seven hundred fifty-five probe sets (representing 571 genes) were differentially expressed at 24 hours after IS in subjects with poor outcome compared to VRFC (FDR-corrected *P* <0.05, FC> |2| (Figure 1a). Of these, 490 probe sets were upregulated and 265 downregulated in subjects with poor outcome (Figure 1a, Table S1C). The 755 probe sets were overrepresented in 58 pathways with seven activated pathways including Regulation of The Epithelial Mesenchymal Transition by Growth Factors Pathway, STAT3 pathway, IL-1 signaling and FGF Signaling (Figure 2c, Table S2C). The B cell receptor signaling and immunoglobulin receptor binding were overrepresented GO terms, including several genes encoding immunoglobulin heavy constant and variably chains, such as genes such as *IGHG3*, *IGHM*, *IGHG1*, *IGHV3-23*, *IGHD*, *IGHA1*, *IGHA2* (FDR < 0.05) (Table S3B). In addition, there was a significant enrichment in neutrophil-specific genes (48/571 genes (8.4%), *P*(overlap) = 1E-07); and in T cell-specific genes (12/571 genes (2.1%), *P*(overlap) = 3E-02 (Figure 3a). Notably, most of the neutrophil-specific genes were up-regulated (41/48), while most of the T cell-specific genes (9/12) were down-regulated in subjects with poor 90-day functional outcome compared to controls.

*24h Gene Expression Associated with Good 90-day Functional Outcome (mRS)*

Fifty probe sets (representing 35 genes) were differentially expressed at 24 hours after IS in subjects with good 90-day outcome compared to VRFC (FDR-corrected *P* <0.05 and FC > |2|) (Figure 1a). Of these, 10 probe sets were upregulated and 40 downregulated in subjects with good outcome (Figure 1a, Table S1C). They were overrepresented in 23 pathways (Figure 2c, Table S2C). In addition, there was a significant enrichment in Erythroblast-specific genes (3/35 genes (8.6%), *P*(overlap) = 1.4E-02) (Figure 3a).

*Differential Expression at 24h Post IS Between Ischemic Stroke Subjects with Poor vs. Good 90-day Functional Outcome (mRS)*

Five hundred fourteen probe sets (representing 363 genes) were differentially expressed between poor vs. good 90-day outcome IS patients with *P* <0.05 and FC> |1.3|. Of these 255 were negatively regulated, and 259 were positively regulated (Figure 1b, Table S1C). They were over-represented in 61 pathways with one being suppressed (ICOS-ICOSL Signaling in T Helper Cells) (Figure 4, Table S2C). Several immune response-related pathways were overrepresented among the 363-gene list, such as Th1 and Th2 Activation Pathway, T Helper Cell Differentiation, Interferon Signaling, and Role of JAK family kinases in IL-6-type Cytokine Signaling (Figure 4, Table S2C). In addition, there was a significant enrichment in neutrophil-specific genes (26/363 genes (7.2%), *P*(overlap) = 1E-03); in T helper-specific, and T cell receptor and signaling-specific genes (2/363 genes (0.6%), *P*(overlap)=4.4E-02 and 8/363 genes (2.2%), *P*(overlap)=2.2E-02, respectively); and in monocyte-specific genes (9/363 genes (2.5%) genes, *P*(overlap) = 6E-03) (Figure 3a). Most of the neutrophil cell-specific genes were up-regulated (21/26), while most of the T cell-specific genes (9/10) were down-regulated in subjects with poor vs. subjects with good 90-day functional outcome.

*24h Gene Expression Correlated with 90-day Outcome (NIHSS)*

Two hundred and one probe sets (representing 147 genes) expressed at 24 hours after IS correlated with 90-day NIHSS (*P*<0.005). Of these, 113 probe sets were negatively correlated and 88 were positively correlated (Figure 1c, Table S1C). The 201 probe sets were over-represented in three pathways (Table S2C and Figure S1). In addition, there was a significant enrichment in T cell-specific genes (5/147 genes (3.4%), *P*(overlap) = 2.5E-03) (Figure 3a). All T cell receptor genes negatively correlated with the 90-day outcome.

**Supplementary Discussion**

Supplementary Discussion (B cells and NK cells).

B cells are part of the adaptive immunity and support neuronal survival, plasticity, recovery, and neurogenesis by producing proteins like neurotrophins to protect neurons [8,9]. Increasing stroke-induced infarct volume and mortality in mice has been shown with lack of B cells [10]. In addition, B cell transfer reduced infarct volumes 3d and 7d after transient middle cerebral artery occlusion (tMCAo) in mice [11]. However, additional studies into long-term role of B cells suggested B cells may contribute to cognitive decline weeks after stroke in mice [12]. Thus, their role is either detrimental or reparative depending on timing, location, and function of B cells/type of the B cells activation recruited into the injured brain [12]. In our study, we observed significant enrichment with B-cell specific-genes in one 5h (90-day NIHSS), and one 24h (90-day NIHSS) negative-beta regression modules (lower expression in worse vs. better NIHSS 90-day outcome), and in the gene lists of Poor Outcome vs. Control at 5h, and Poor vs. Good Outcome at 5h and 24h mRS outcome. Most all B cell-specific genes were down-regulated in poor outcome. On the other hand, in modules different from the ones enriched with B cell-specific genes, the B Cell Receptor Signaling was significantly activated in subjects in poor vs. good 90-day mRS outcome in one 3h module, two 5h modules and one 24h module. All four modules were positive-beta regression modules for 90-day mRS (higher eigengene expression in poor vs. good outcome). In addition, B Cell Receptor Signaling was significantly suppressed in one 24h module that negatively associated with 90-day mRS, which was also enriched in B-cell specific genes. In that module, the more suppressed the B Cell Receptor Signaling Pathway, the higher the mRS, the worse the 90-day mRS outcome. Thus, our data underscores the complex involvement of B cells in the early peripheral B cell immune response to ischemic stroke. The identified genes may be potential therapeutic targets.

NK cells are a critical component of the innate immune system. They infiltrate the injured brain following stroke and are detected in the peri-infarct region [13]. They have been associated with inflammation and infections after stroke, and with exacerbation of brain infarction, BBB damage and infarct size [13]. Recruitment of NK cells by ischemic neurons has been shown to accelerate brain infarction [14]. In addition, decreased NK cell counts in peripheral blood in days one, three and seven following IS have also been reported [15]. In our data, NK-specific genes were significantly enriched in one 3h module, one 5h module (and its hubs), and one 24h module – all with negative beta regression coefficient with 90-day mRS (the lower the expression, the higher the 90-day mRS, the worse the 90-day outcome). Additional studies will need to further dissect the mechanisms and function of NK cells in the peripheral immune system and in the injured brain post stroke and how this affects outcome.
