## Supplemental Figure 1 and 2 for "Peripheral Blood Gene Expression at 3 to 24 Hours Correlates with and Predicts 90-Day Outcome Following Human Ischemic Stroke"

### Fig. S1 Pathway Enrichment in Genes Expression Correlates with 90-day Outcome (NIHSS)

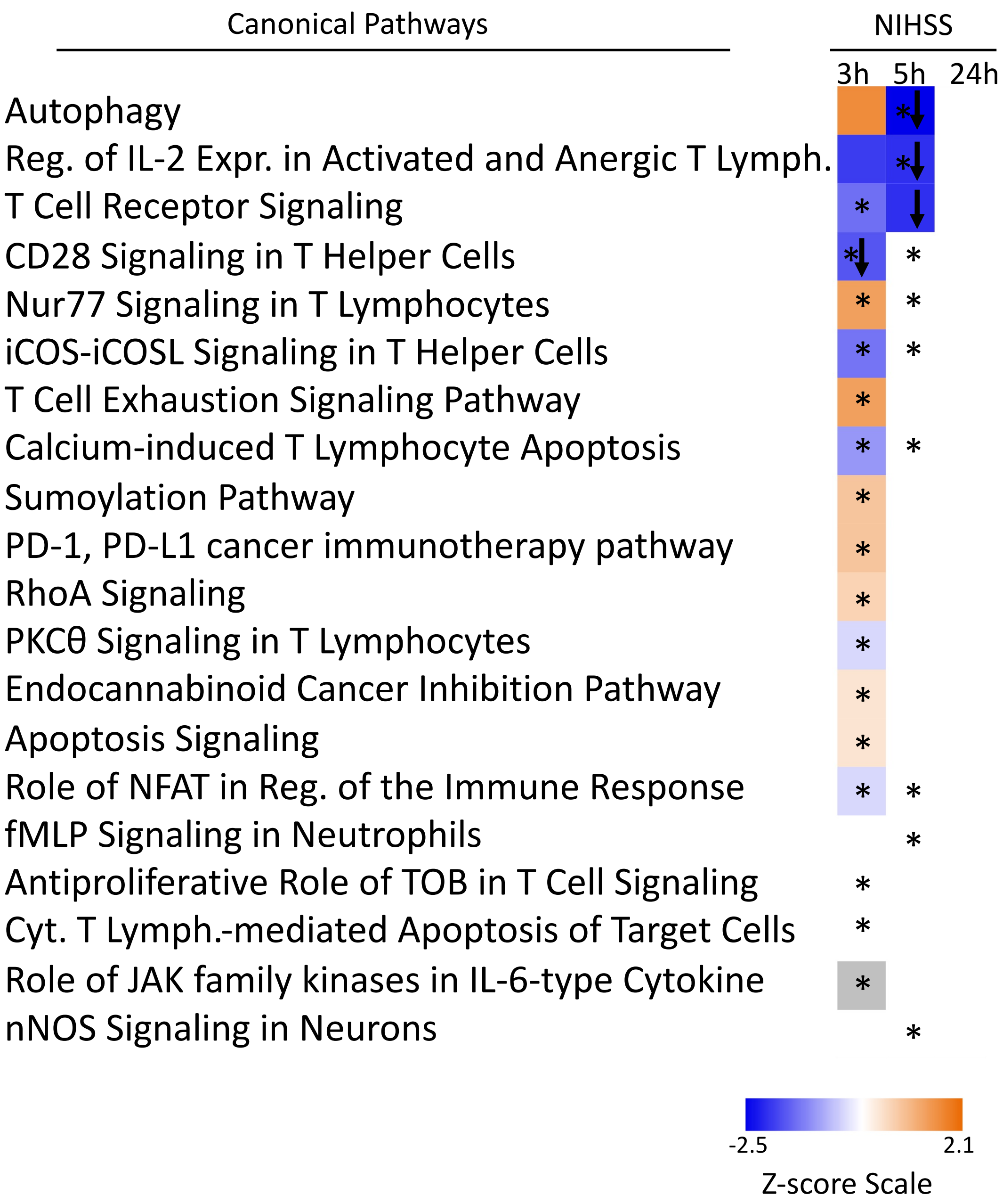

Fig. S2

5hCyan- Upregulated in Poor Outcome vs. Good 90-Day IS Outcome

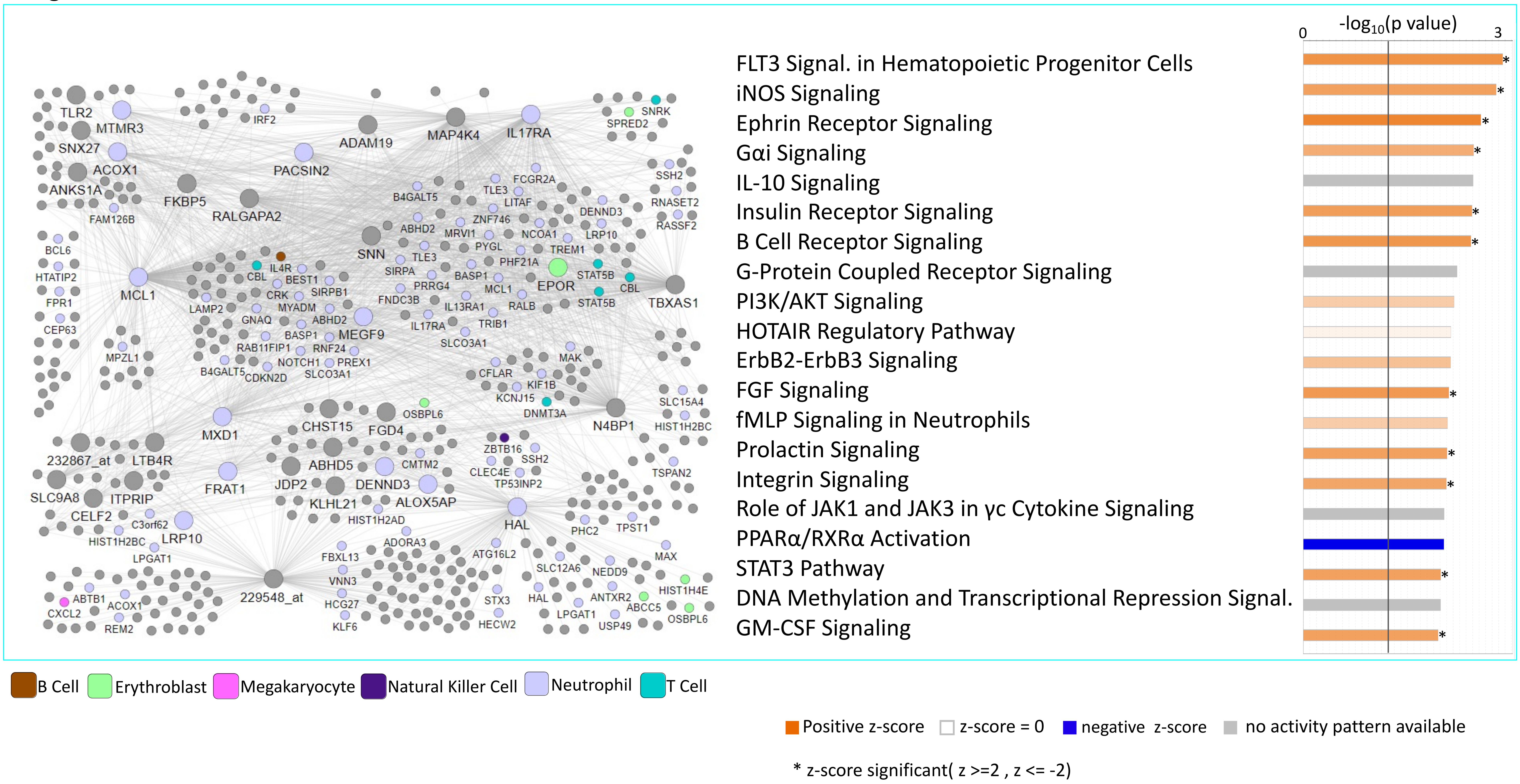
